# Effect of Intensity Discretization on Radiomic Features in En Face EDI-OCT of Choroidal Nevi

**DOI:** 10.64898/2026.11.01.26367028

**Authors:** Marta Vila Franca, João P. Nobre, Duarte Saraiva, Helena Rico Pereira, Hugo Duarte Ferreira, Marco Dutra Medeiros, Maria da Luz Cachulo

## Abstract

**Background/Objectives:** Intensity discretization influences radiomic measurements, but its effects in optical coherence tomography (OCT) of choroidal lesions remain insufficiently characterized. We evaluated fixed bin width (FBW) and fixed bin number (FBN) strategies with different parameters to inform discretization choices for en face enhanced-depth imaging OCT (EDI-OCT) of stable choroidal nevi.

**Methods:** Ninety-three original-domain radiomic features were extracted from 64 eyes across 11 discretization settings (FBW 2–64; FBN 8–64). We assessed feature values, eye rankings, observer reproducibility, slab-depth dependence, redundancy, and feature-set overlap. Primary absolute-agreement intraclass correlation coefficients (ICCs) used per-eye feature values averaged across six numeric slabs. Histogram entropy and an approximate co-occurrence sampling-density proxy characterized the discretized intensity representation.

**Results:** Seventy-seven features depended on discretization. Median occupied gray levels decreased with increasing bin width from 54 at FBW 2 to five at FBW 25. Eye rankings were similar between neighboring settings (median Spearman *ρ*, 0.97–0.99), but less concordant in selected FBW–FBN comparisons. Median observer ICCs ranged from 0.84 to 0.91, without clear evidence of differences between settings. Feature sets overlapped less after redundancy reduction. FBW 4 and FBN 32 provided intermediate gray-level resolution and sampling-density proxies, retaining at least 16 occupied gray levels in approximately 99% and 100% of observations, respectively.

**Conclusions:** Intensity discretization substantially alters radiomic texture values, eye rankings, and feature selection. Observer reproducibility alone is insufficient to guide discretization choice. Intensity representation, gray-level compression, and slab depth should also be considered. FBW 4 and FBN 32 emerged as exploratory candidate settings within the evaluated pipeline. External validation and assessment of clinical utility remain necessary.

## 1 Introduction

Choroidal nevus (CN) is the most common intraocular tumor. Histopathologic[1, 2] evidence suggests that melanocytic choroidal tumors comprise a continuous spectrum ranging from benign-appearing melanocytes to atypical-appearing melanocytes (Spindle A) aggregated to form solid lesions,[3] with an overlap between the two, making their differentiation a significant challenge.[4, 5] Although most nevi remain stable, a small subset undergoes malignant transformation into melanoma, therefore early risk stratification is critical[6, 7] and relies on multimodal imaging[8], including optical coherence tomography (OCT)[9, 10]. The imaging risk factors [8, 11, 12] currently used, such as lesion growth, presence of subretinal fluid, or orange pigment, are largely qualitative and observer dependent. Consequently, some pigmented choroidal lesions remain clinically indeterminate[4, 13]. Therefore, objective quantitative biomarkers of the internal structure of CN extracted from routine OCT images are needed to characterize these lesions reproducibly and to support early identification of lesions at risk.

Radiomics is a promising approach[14–18] for obtaining quantitative imaging biomarkers. It converts medical images into quantitative data based on the assumption that they contain information related to tissue biology that is imperceptible to the human eye[18–20]. In handcrafted radiomics, predefined features are extracted from a segmented region of interest (ROI) by applying mathematical formulas to the pixel intensities. These handcrafted radiomic features (HRF) include first-order, shape, and texture features, with the latter quantifying the spatial arrangement of pixel intensities and, by extension, tissue heterogeneity[17, 21–23]. Unlike deep learning approaches, in which the learned parameters vary between model instances and result in a “black box”, HRF are transparent and interpretable, do not require model training, and can be applied to relatively small cohorts[24]. Radiomics is well established in oncology and radiology and is increasingly being applied in ophthalmology[25].

The reliability of HRF depends on several factors, including image acquisition, reconstruction, preprocessing and ROI segmentation[14, 26, 27]. Texture analysis is particularly dependent on intensity discretization[28, 29], as texture features are computed on matrices generated after grouping gray levels into bins. This grouping can be performed using two different approaches. Absolute discretization via fixed bin width (FBW) uses bins of a constant size, and the resulting number of bins depends on the intensity range of the ROI. In contrast, relative discretization via fixed bin number (FBN) scales bin widths to divide the total intensity range into a fixed number of intervals. Both the discretization method and its parameters can alter the resulting matrices and, consequently, feature values, redundancy, and reproducibility[27]. Too few bins discard subtle intensity differences, whereas too many generate sparse matrices that amplify noise. In other imaging modalities, such as computed tomography (CT), magnetic resonance imaging (MRI) and positron emission tomography (PET), recommendations for binning strategies have been proposed, commonly expressed as a number of gray levels. Practical recommendations in radiomics studies in PET have suggested choosing a bin width that yields 16-128 or 30-130 bins[30, 31]. In OCT however, image intensities are device-dependent and not standardized or calibrated across systems, and therefore recommendations from other imaging modalities cannot be directly transposed to OCT. To the best of our knowledge, the effect of different intensity discretization strategies on radiomic features extracted from OCT images of choroidal lesions has not yet been systematically investigated and has only begun to be explored in dermatological OCT[32].

The choice of intensity discretization strategy is important as it may determine which features can be reliably extracted to characterize choroidal lesions and therefore the feature set available for building subsequent radiomic signatures or predictive models. In this study, we evaluated the effect of intensity discretization on the reproducibility and stability of HRF extracted from en face enhanced-depth imaging OCT (EDI-OCT) images of stable CN, aiming to identify settings that balance information retention, matrix density, and reproducibility for subsequent analyses.

## 2 Objectives

The primary aim of this study was to systematically evaluate the effect of intensity discretization on the reproducibility and stability of HRF extracted from en face EDI-OCT images of stable CN and to identify settings that balance information retention, matrix density and reproducibility for these lesions.

Specifically, we compared fixed bin width (FBW) and fixed bin number (FBN) strategies to assess: their effect on feature values and on the ranking of eyes by feature value; intra- and inter-observer reproducibility, using intraclass correlation coefficients (ICC); the dependence of feature values on choroidal slab depth; feature redundancy and the composition of the resulting non-redundant feature sets; and the trade-off between information retention, texture matrix density and the number of gray levels relative to the commonly advised range of 16 to 128, in order to identify settings that balance them.

## 3 Materials and Methods

### 3.1 Study design and participants

A cross-sectional study was conducted at two centers, the Departments of Ophthalmology of ULS of Santa Maria and ULS of São José, between June and October 2024. The study was approved by the institutional Bioethics Committees and conducted in accordance with the Declaration of Helsinki. Written informed consent was obtained from all participants.

Adults with a known choroidal nevus in the posterior pole, largest basal diameter < 5 mm, thickness < 2 mm, at least 3 years of documented follow-up, and no documented growth were eligible. Exclusion criteria included pregnancy, active intraocular inflammation or infection, retinal or choroidal disease affecting image analysis, refractive error > 6 diopters, prior intraocular surgery other than cataract surgery, and poor OCT image quality (signal strength < 25 dB). All participants underwent dilated fundus examination and macular OCT and OCT angiography. Sixty-four participants fulfilled the eligibility criteria and completed the imaging protocol.

### 3.2 Image acquisition and ROI segmentation

OCT images were acquired after pupillary dilation using enhanced-depth imaging on a Spectralis OCT system (Heidelberg Engineering, Heidelberg, Germany). A 20° × 20° high-resolution volume centered on the lesion was acquired with 97 B-scans, automatic real-time averaging, and active eye tracking. En face reconstructions and image export were performed using Heidelberg Eye Explorer (HEYEX), version 2.6.4. Six contiguous 20 *μ*m-thick en face slabs were generated, with anterior boundaries positioned 20, 40, 60, 80, 100, and 120 *μ*m posterior to the lower boundary of the retinal pigment epithelium (RPE)/Bruch’s membrane complex. Slab labels indicate their anterior boundary; thus, the six slabs covered depth intervals of 20–40, 40–60, 60–80, 80–100, 100–120, and 120–140 *μ*m, respectively. An additional TOTAL slab extended from the anterior border of the nevus to the depth at which the lesion signal began to fade. En face images were generated using maximum-intensity projection, and anatomical coverage was assessed by visual inspection. Images were exported as RGB tagged image file format (TIFF) files followed by grayscale conversion. Brightness and contrast settings were left unchanged across images during export.

ROIs were manually segmented on the Total slab using a custom Python application (RadiomicsOCT) and propagated to the six depth-specific slabs, with manual adjustment when required. Three independent segmentation rounds were performed: two by the same observer separated by 2 weeks and one by a second observer. Radiomic extraction was repeated for all discretization settings using identical images and masks.

### 3.3 Radiomic features and discretization

Ninety-three radiomic features were extracted using PyRadiomics[33], including 18 firstorder features and 75 texture features from gray-level co-occurrence matrix (GLCM), gray-level run-length matrix (GLRLM), gray-level size-zone matrix (GLSZM), gray-level dependence matrix (GLDM), and neighboring gray-tone difference matrix (NGTDM) families. Shape and filtered-image features were excluded. Full extraction parameters are provided in the Supplementary Methods.

Eleven discretization settings were compared: seven fixed bin width settings (FBW 2, 4, 8, 16, 25, 32, and 64) and four fixed bin number settings (FBN 8, 16, 32, and 64). FBW used fixed-width intensity intervals and FBN divided each ROI intensity range into a pre-defined number of bins. We assessed the number of occupied gray levels, histogram entropy, and approximate GLCM density, defined as the ROI pixel count divided by the square of the number of occupied gray levels. This density measure was a proxy rather than actual matrix occupancy. Histogram entropy was used as a surrogate for the information content of the discretized intensity representation. Higher entropy indicates greater intensity diversity but does not necessarily imply greater biological or diagnostic information. Detailed quantization definitions and extraction details are provided in the Supplementary Methods.

### 3.4 Statistical analysis

Discretization effects were assessed using fold-changes relative to FBW 2 and Spearman rank correlations. Intra and inter-observer reproducibility was evaluated using absoluteagreement intraclass correlation coefficients (ICCs)[34], with the six numeric slabs summarized in the primary analysis and individual slabs and the TOTAL slab assessed separately. Features were considered reproducible when ICC was ≥ 0.80 in both the intraobserver and primary inter-observer comparisons.

Depth-related variation was assessed using the Friedman test and Kendall’s W and monotonic trends, with Benjamini-Hochberg correction[35]. Reproducible features were reduced using consensus absolute Spearman correlations |*ρ*| ≥ 0.90, retaining one representative per correlation group. Feature grouping and final set overlap were compared using the adjusted Rand and Jaccard indices, respectively.

## 4 Results

### 4.1 Study population

A total of 302 patients were initially identified for potential inclusion. Following medical record review, 203 patients were excluded for not meeting the predefined inclusion criteria. An additional 35 patients were excluded after clinical evaluation. Ultimately, 64 patients (mean age 71.9 ±10.4 years; 56.3% female) were included. All patients were white. None of the patients reported new symptoms at the time of the study.

### 4.2 Choroidal nevus characteristics

The clinical and multimodal imaging characteristics of the choroidal nevi are summarized in Table 1. The mean largest basal diameter was 2714 ± 814.7 *μ*m, and the mean nevus thickness, measured by OCT, was 316.4 ± 140.4 *μ*m. Subretinal fluid (SRF) was present in 7 (10.9%) nevus-affected eyes, while drusen were observed in 26 (40.6%). Most lesions were flat (*n* = 45; 70.3%), 18 (28.1%) were dome-shaped and 1 (1.6%) was almond-shaped. Among dome-shaped lesions, the mean dome height was 173.1 ± 140.1 *μ*m.

**Table 1.** Clinical and multimodal imaging characteristics of choroidal nevi in 64 study eyes.

| **Characteristic** | *N* **(%) or Mean** ± **SD** |
|---|---|
| **Nevus location** |  |
| Temporal-superior | 15 (23.4%) |
| Temporal-inferior | 15 (23.4%) |
| Macular | 14 (21.9%) |
| Juxtapapillary | 10 (15.6%) |
| Nasal-superior | 3 (4.7%) |
| Nasal-inferior | 7 (10.9%) |
| **Nevus size (*μ*m)** |  |
| Largest basal diameter | 2714.0 ± 814.7 |
| Thickness | 316.4 ± 140.4 |
| **Nevus configuration** |  |
| Flat | 45 (70.3%) |
| Dome-shaped | 18 (28.1%) |
| Almond-shaped | 1 (1.6%) |
| Mean dome height (*μ*m)\* | 173.1 ± 140.1 |
| **Autofluorescence** |  |
| Isoautofluorescent | 52 (81.3%) |
| Hyperautofluorescent | 12 (18.8%) |
| **Intrinsic hyperreflectivity** | 62 (96.9%) |
| **Reflectivity on OCT** |  |
| Hyperreflective | 54 (84.4%) |
| Isoreflective | 9 (14.1%) |
| Hyporeflective | 1 (1.6%) |
| **Associated features** |  |
| Drusen | 26 (40.6%) |
| Subretinal fluid | 7 (10.9%) |
| CNV type 1 | 3 (4.7%) |

<sup>1</sup> Dome height calculated only for dome-shaped lesions (*n* = 18). Values are *n* (%) or mean ± standard deviation; OCT = optical coherence tomography; CNV = choroidal neovascularization; SD = standard deviation.

### 4.3 ROI size and segmentation agreement

ROI size varied substantially between eyes with a median of 42,970 pixels (interquartile range [IQR], 27,042-61,216) in observer 1, session 1. ROI-size agreement was high for the intra-observer comparison (ICC: 0.948; 95% confidence interval [CI], 0.883–0.978) and the primary inter-observer comparison (ICC: 0.948; 95% CI, 0.867–0.974).

### 4.4 Properties of the discretization settings

We evaluated 11 discretization settings (Figure 1, Table 2 and Supplementary Figure S1). With FBN, the nominal bin count was fixed by design, while the number of occupied levels could be slightly lower, whereas with FBW it decreased as bin width increased. The median number of gray levels decreased from 54 at FBW 2 to 14 at FBW 8, 5 at FBW 25 and 2 at FBW 64. At FBW 25, 98% of the ROIs had fewer than 8 gray levels, compared with 0% at FBW 4. Discretized histogram entropy increased with finer discretization. FBW 2 and FBN 64 yielded the highest histogram entropy (4.89 and 5.12, respectively), whereas FBW 25 and FBW 64 yielded lower entropy values (1.44 and 0.68 bits). In contrast, coarse settings yielded higher approximate GLCM densities, with a median of 215 pixels per cell at FBW 8 and 1,810 at FBW 25, compared with 14.4 at FBW 2 and 10.6 at FBN 64.

**Figure 1.**
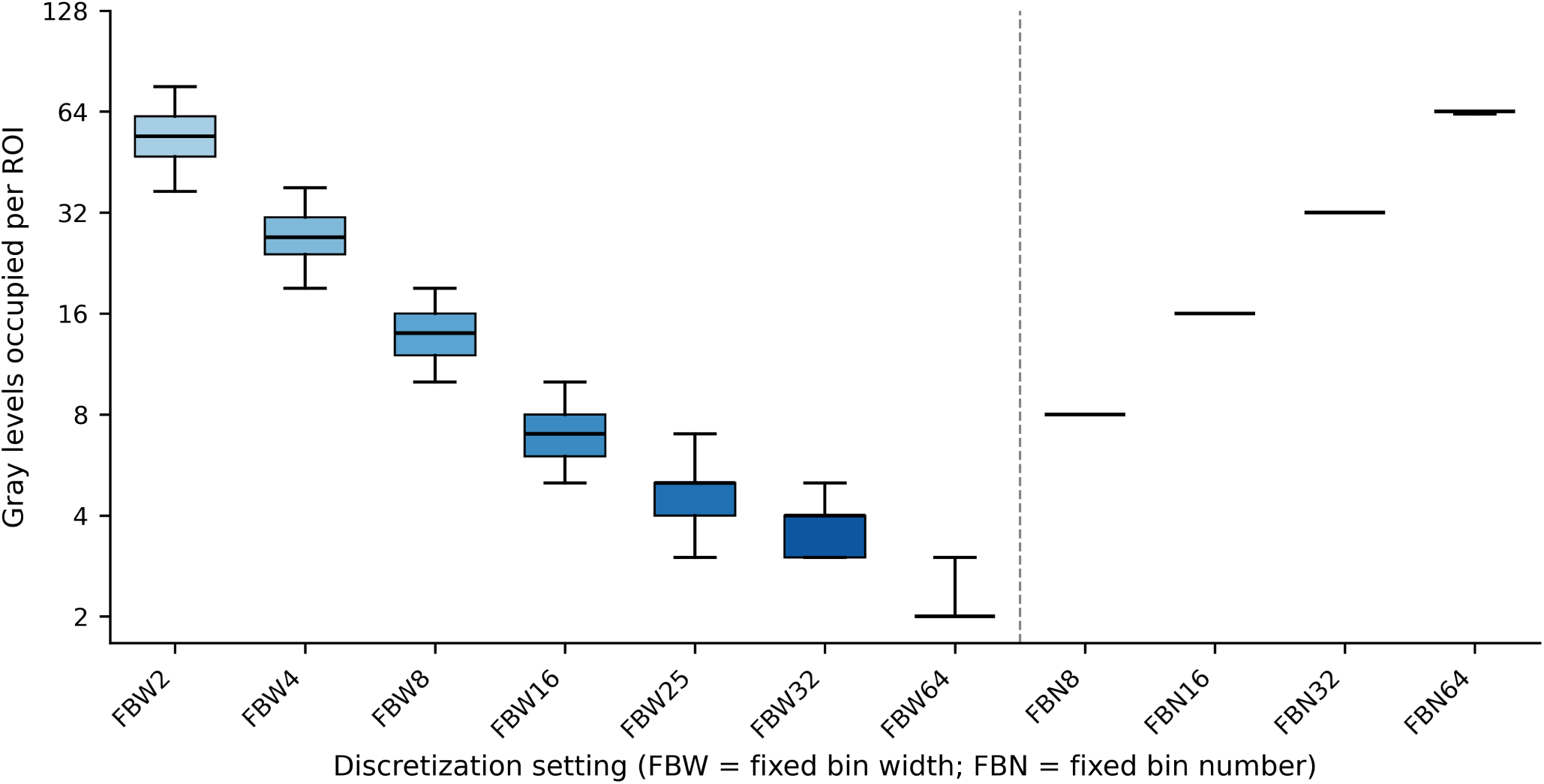
Occupied gray levels across discretization settings. The distributions summarize 448 ROI-slab observations from 64 eyes and seven slab views, using observer 1, session 1. Boxes represent the interquartile range, central lines the median and whiskers the 5^th^-95^th^ percentiles. The vertical dashed line separates fixed bin width (FBW) from fixed bin number (FBN) settings. The vertical axis is logarithmic. ROI, region of interest.

**Table 2.** Quantization properties of the 11 discretization settings.

| **Setting** | **Occupied gray levels, median [P5–P95]** | **Observations with <8 / <16 occupied levels, %** | **Approximate GLCM density\*, median** | **Entropy (bits), median [IQR]** | **Effective gray levels†** |
|---|---|---|---|---|---|
| FBW 2 | 54 [36–77] | 0 / 0 | 14.4 | 4.89 [4.65–5.16] | 29.6 |
| FBW 4 | 27 [18–39] | 0 / 1 | 56.4 | 3.90 [3.65–4.17] | 14.9 |
| FBW 8 | 14 [9–20] | 1 / 71 | 215.1 | 2.91 [2.67–3.18] | 7.5 |
| FBW 16 | 7 [5–10] | 60 / 100 | 824.3 | 1.98 [1.76–2.24] | 3.9 |
| FBW 25 | 5 [3–7] | 98 / 100 | 1,810.1 | 1.44 [1.25–1.68] | 2.7 |
| FBW 32 | 4 [3–5] | 100 / 100 | 2,917.6 | 1.17 [1.02–1.40] | 2.2 |
| FBW 64 | 2 [2–3] | 100 / 100 | 8,868.4 | 0.68 [0.39–0.93] | 1.6 |
| FBN 8 | 8 [8–8] | 0 / 100 | 671.4 | 2.23 [2.10–2.37] | 4.7 |
| FBN 16 | 16 [16–16] | 0 / 0 | 167.9 | 3.18 [3.05–3.32] | 9.1 |
| FBN 32 | 32 [32–32] | 0 / 0 | 42.0 | 4.16 [4.02–4.30] | 17.9 |
| FBN 64 | 64 [63–64] | 0 / 0 | 10.6 | 5.12 [4.97–5.26] | 34.8 |

Summaries are based on 448 ROI-slab observations (64 eyes x 7 slab views) using observer 1, session 1. Occupied levels are bins containing at least one ROI pixel. \*Approximate GLCM density: ROI count divided by the square of occupied gray levels; this is not a measure of actual matrix occupancy. † Effective gray levels: 2 raised to the median entropy. FBW, fixed bin width; FBN, fixed bin number; ROI, region of interest; GLCM, gray-level co-occurrence matrix; P5–P95, 5th–95th percentiles; IQR, interquartile range.

### 4.5 Effect on feature values and rankings

Of the 93 features, 16 first-order features were independent of discretization, whereas the remaining 77 features, mainly texture features, varied across settings. Feature values changed progressively as bin width increased, with substantial changes already evident at FBW 25 (Figure 2). At this setting, median changes ranged from 8-fold for GLCM features to 160-fold for NGTDM features, and most texture features changed by more than two-fold. At FBN 64, typical fold-changes relative to FBW 2 ranged from 1.2 to 1.9 across feature families. Neighboring settings produced similar rankings of eyes. The median Spearman correlation was 0.99 for FBW 2 vs FBW 4 and 0.97 for FBN 32 vs FBN 64, but decreased when more distant settings or different discretization methods were compared (Figure 3). Feature values from FBW settings were also more strongly related to the ROI intensity range than those from FBN settings, whereas neither approach showed a strong association with mean ROI intensity.

**Figure 2.**
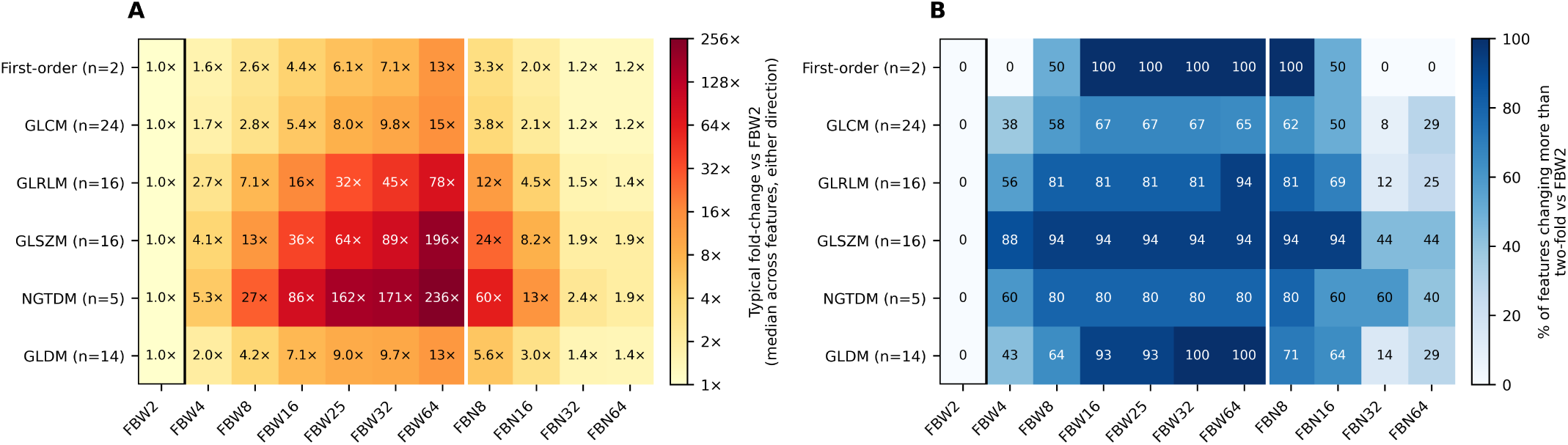
Changes in radiomic feature values relative to FBW 2. (A) Typical fold-change by feature family, summarized without regard to direction as 2 raised to the median absolute log2 fold-change. (B) Percentage of features with a change greater than two-fold. Analysis included the 77 discretization-dependent features in the TOTAL slab from observer 1, session 1, across 64 eyes. The first-order family comprises entropy and uniformity; the 16 discretization-independent first-order features are excluded. Ratios were excluded when the reference median was zero or the sign changed. GLCM, gray-level co-occurrence matrix; GLRLM, gray-level run-length matrix; GLSZM, gray-level size-zone matrix; NGTDM, neighboring gray-tone difference matrix; GLDM, gray-level dependence matrix; FBW, fixed bin width; FBN, fixed bin number.

**Figure 3.**
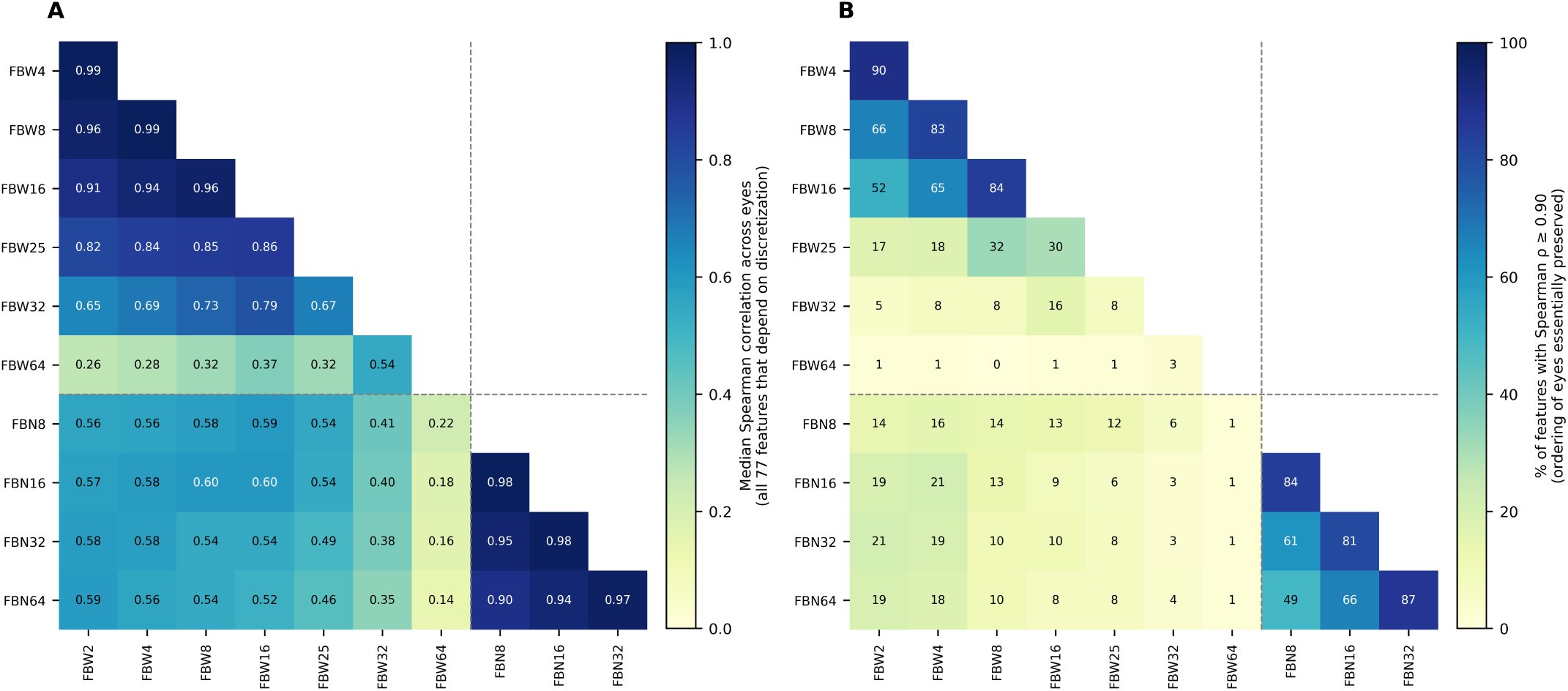
Preservation of eye rankings between discretization settings. For each of the 77 discretization-dependent features, Spearman correlations were calculated across 64 eyes for each pair of settings using the TOTAL slab from observer 1, session 1. (A) Median correlation across features. (B) Percentage of features with Spearman’s *ρ* ≥0.90. Dashed lines separate fixed bin width (FBW) and fixed bin number (FBN) settings.

### 4.6 Feature reproducibility

Median ICCs were high across all discretization settings, ranging from 0.87 to 0.91 for intra-observer and from 0.84 to 0.90 for inter-observer reproducibility (Figure 4). Differences between settings were small compared with the variability between individual features, and the paired bootstrap comparisons did not provide clear evidence of differences between settings, though this does not establish equivalence. FBN settings had a slightly higher proportion of features with ICCs ≥ 0.75 than FBW settings, although the confidence intervals overlapped. At FBW 64, where ROIs typically contained only two gray levels, NGTDM features showed poor reproducibility (ICC approximately 0.2-0.5), making this the only setting with poorly reproducible features.

**Figure 4.**
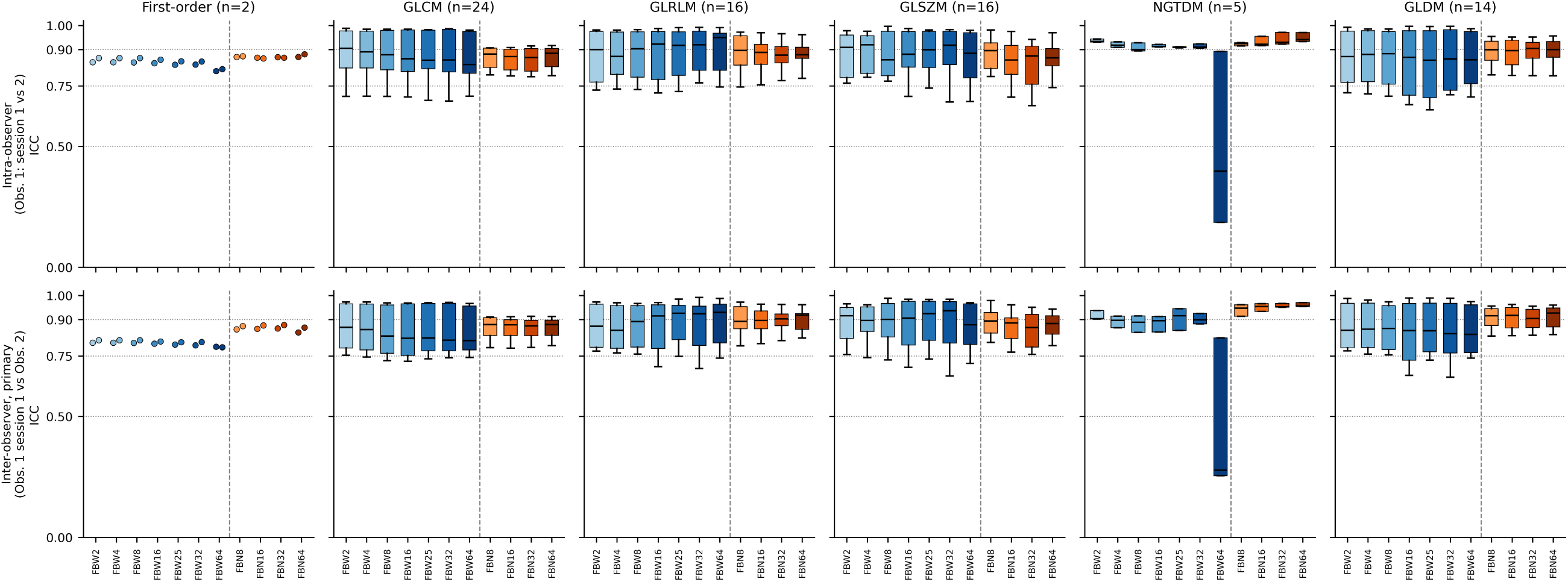
Observer reproducibility across feature families and discretization settings. Primary depth-summary ICCs are shown for the intra-observer comparison (observer 1, session 1 versus session 2; top row) and primary inter-observer comparison (observer 1, session 1 versus observer 2; bottom row). The 77 discretization-dependent features are grouped by family. Boxes show the interquartile range and the central lines the median; first-order entropy and uniformity are shown as individual points. Horizontal reference lines indicate ICC values of 0.50, 0.75 and 0.90. Vertical dashed lines separate FBW and FBN. ICC, intraclass correlation coefficient; FBW, fixed bin width; FBN, fixed bin number; GLCM, gray-level co-occurrence matrix; GLRLM, gray-level run-length matrix; GLSZM, gray-level size-zone matrix; NGTDM, neighboring gray-tone difference matrix; GLDM, gray-level dependence matrix.

**Figure 5.**
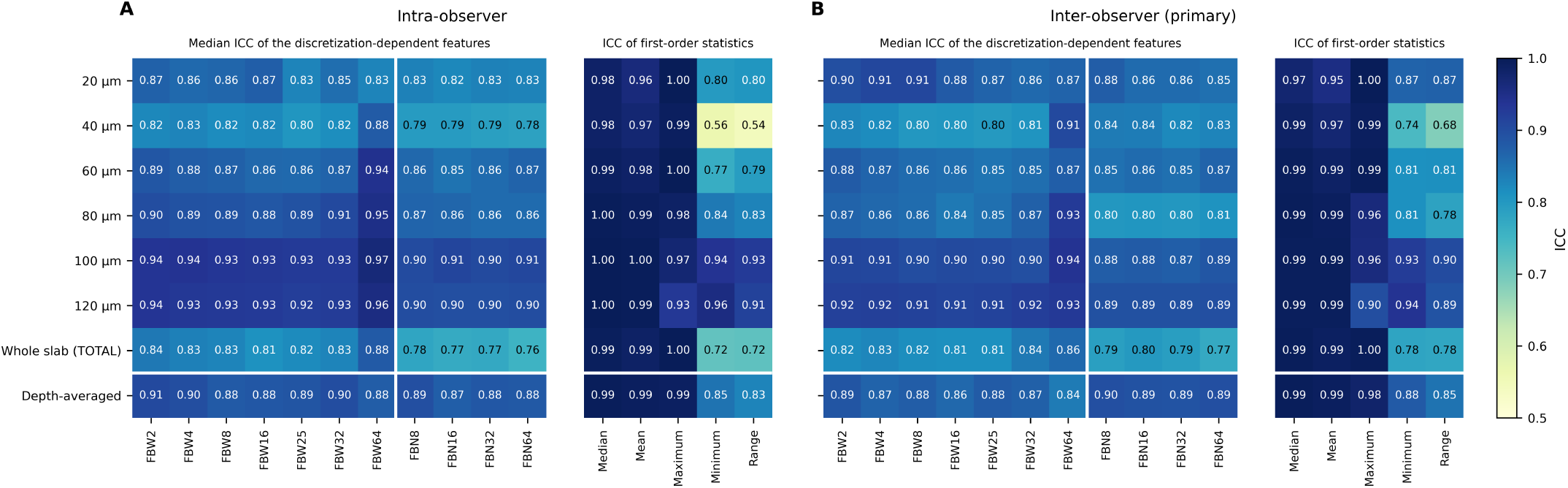
Reproducibility by slab depth and discretization setting. (A) Intra-observer and (B) primary inter-observer ICCs. Heatmaps show median ICCs across the 77 discretization-dependent features at each depth, for the TOTAL slab and for the primary depth-summary analysis. Adjacent reference columns show ICCs for five first-order intensity statistics that do not depend on discretization. Cell labels display ICC values; colors indicate their magnitude. Vertical dashed lines separate FBW and FBN settings. ICC, intraclass correlation coefficient; FBW, fixed bin width; FBN, fixed bin number.

Reproducibility was lower for the TOTAL slab, with median ICCs ranging from 0.76 to 0.88. FBW 4 and FBN 32 showed reproducibility within the range observed across all settings (Table 3). ICCs and concordance correlation coefficients showed similar results.

**Table 3.** Summary of quantization, reproducibility and feature relationship for representative discretization settings.

| **Setting** | **Occupied gray levels, median** | **Entropy (bits)** | **Median depth-summary ICC (intra / inter)** | **Depth effect (median W)** | **Correlation groups among 77 features** | **Ranking agreement vs FBW 2 / vs FBN 64** |
|---|---|---|---|---|---|---|
| FBW 2 | 54 | 4.89 | 0.91 / 0.89 | 0.31 | 30 | – / 0.59 |
| FBW 4 | 27 | 3.90 | 0.90 / 0.87 | 0.31 | 32 | 0.99 / 0.56 |
| FBW 8 | 14 | 2.91 | 0.88 / 0.88 | 0.29 | 32 | 0.96 / 0.54 |
| FBW 25 | 5 | 1.44 | 0.89 / 0.88 | 0.22 | 37 | 0.82 / 0.46 |
| FBN 32 | 32 | 4.16 | 0.88 / 0.89 | 0.25 | 33 | 0.58 / 0.97 |
| FBN 64 | 64 | 5.12 | 0.88 / 0.89 | 0.28 | 28 | 0.59 / – |

Gray levels and entropy are medians across 448 ROI-slab observations. Median ICCs summarize 77 discretization-dependent features for the primary depth-summary analysis: intra/inter denotes intra-observer and primary inter-observer comparisons. W is the median Kendall’s W across features. Correlation groups are defined using consensus \|*ρ*\| ≥ 0.90 threshold and are not counts of the final reproducible feature sets. Ranking agreement is the median, across the 77 features of Spearman correlation calculated across 64 eyes in the TOTAL slab from Observer 1, Session 1. ICC, intraclass correlation coefficient, FBW; fixed bin width; FBN, fixed bin number.

### 4.7 Variation with slab depth

Reproducibility varied more across slab depth than across discretization settings (Figure 5A). Median intra-observer ICCs were lowest at 40 *μ*m (0.78-0.83) and highest at 100-120 *μ*m (0.90-0.94). At 40 *μ*m, intensity range and minimum were the least reproducible first-order features (ICC 0.54 and 0.56, respectively). FBN ICCs were generally slightly lower than those for FBW. Although FBW 64 showed the highest ICCs in deeper slabs, it retained very little information.

Image characteristics also changed with depth. Mean ROI intensity decreased by 43% at 120 *μ*m relative to the 20 *μ*m slab. Entropy decreased with depth under FBW, particularly at wider bin widths, but remained relatively stable under FBN 64. Contrast decreased across all settings, by 28%-46% at 120 *μ*m. Almost all features varied significantly with depth (94-100% across settings), although the magnitude of these changes varied. Median Kendall’s W ranged from 0.09 to 0.31 for FBW and from 0.24 to 0.28 for FBN. Depth-related trends were monotonic for most features, particularly under FBW. Depth-profiles and effect-size summaries are shown in Supplementary Figures S4 and S5.

### 4.8 Redundancy and reproducible feature selection

Among the 77 discretization-dependent features, the number of correlation groups, defined using a consensus |*ρ*| ≥ 0.90 threshold, ranged from 28 to 41 across the 11 settings (Supplementary Figure S6). The number of groups was negatively correlated with the number of occupied gray levels, Spearman *ρ* = −0.92. FBW 4, FBW 8 and FBN 32 yielded 32, 32 and 33 groups, respectively. Feature grouping was more similar between settings with comparable numbers of gray levels (adjusted Rand index 0.67-0.83) than between settings with different numbers (0.22-0.37).

The number of features with *ICC* ≥ 0.80 for both intra- and inter-observer comparisons ranged from 56 to 65 for FBW and from 72 to 78 for FBN. After redundancy reduction (Figure 6), the final sets contained 24-31 features for FBW and 31-34 for FBN. Overlap between settings and feature-selection frequencies are illustrated in Supplementary Figure S7. When restricted to discretization-dependent features, the median Jaccard index was 0.74 before redundancy reduction and 0.40 after reduction. For neighboring discretization settings, the index ranged from 0.89 to 0.91 before reduction and from 0.54 to 0.74 after reduction.

**Figure 6.**
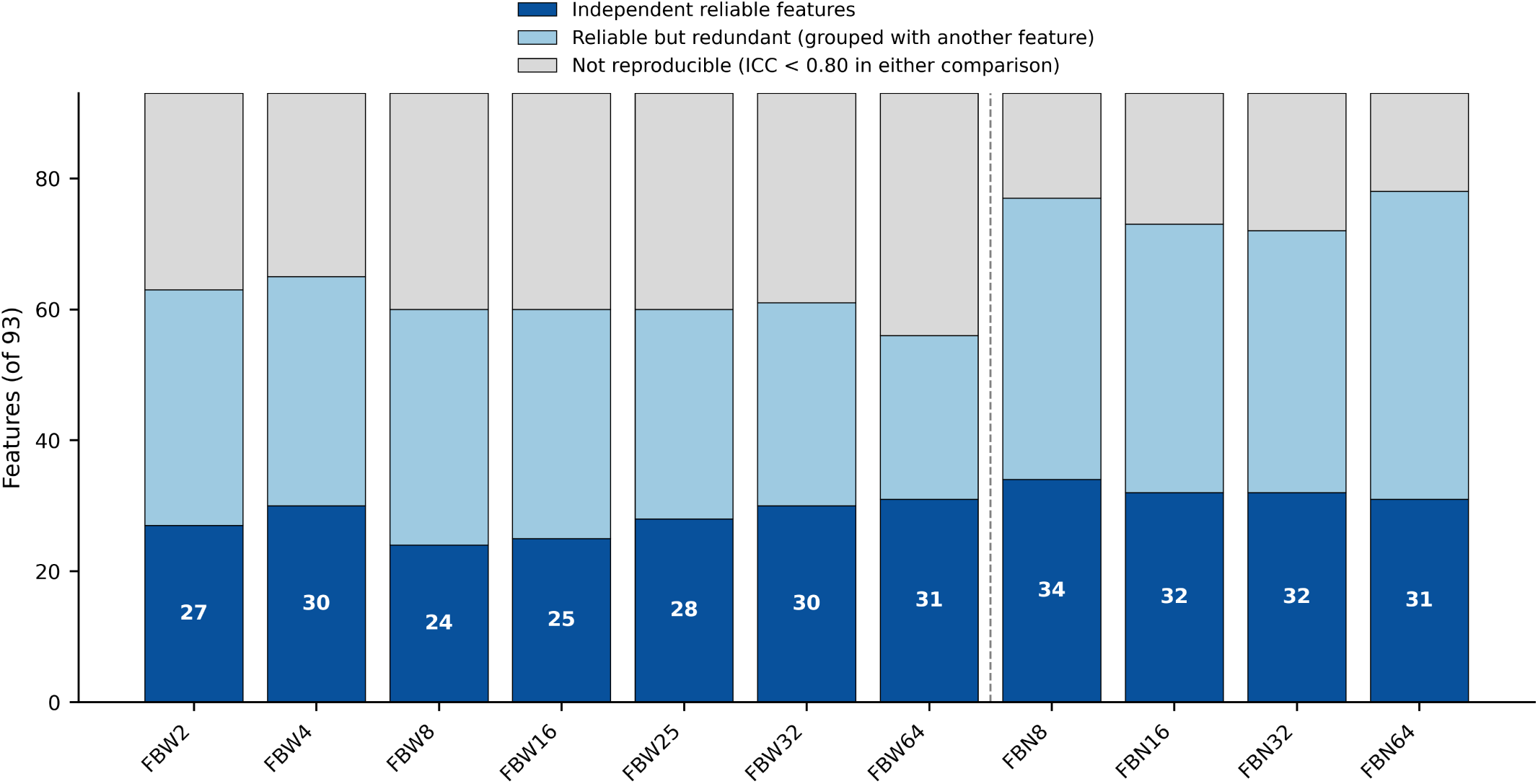
Reproducibility and redundancy reduction across discretization settings. Each bar represents all 93 extracted features. Dark blue indicates the final representative features retained after reproducibility screening and redundancy reduction; light blue indicates reproducible features grouped with another retained feature; gray indicates features not meeting the reproducibility criterion. Reproducibility required ICC ≥0.80 in both the intra-observer and primary inter-observer comparisons. Correlated features were grouped using a consensus |*ρ*| ≥0.90 threshold, and one representative was retained per group. White labels indicate final feature counts. The vertical dashed line separates FBW and FBN settings. ICC, intraclass correlation coefficient; FBW, fixed bin width; FBN, fixed bin number.

The final feature sets contained 5-7 first-order features, which were independent of discretization, together with texture features, most frequently from the GLSZM family.

### 4.9 Information, matrix density and choice of setting

Across the 11 discretization settings, histogram entropy increased as approximate matrix density decreased (Figure 7). FBN 64 retained 5.12 bits (95% CI, 5.08-5.16), compared with 4.89 bits (95% CI, 4.83-4.97) for FBW 2, with median matrix densities of 10.6 and 14.4 pixels per cell, respectively.

**Figure 7.**
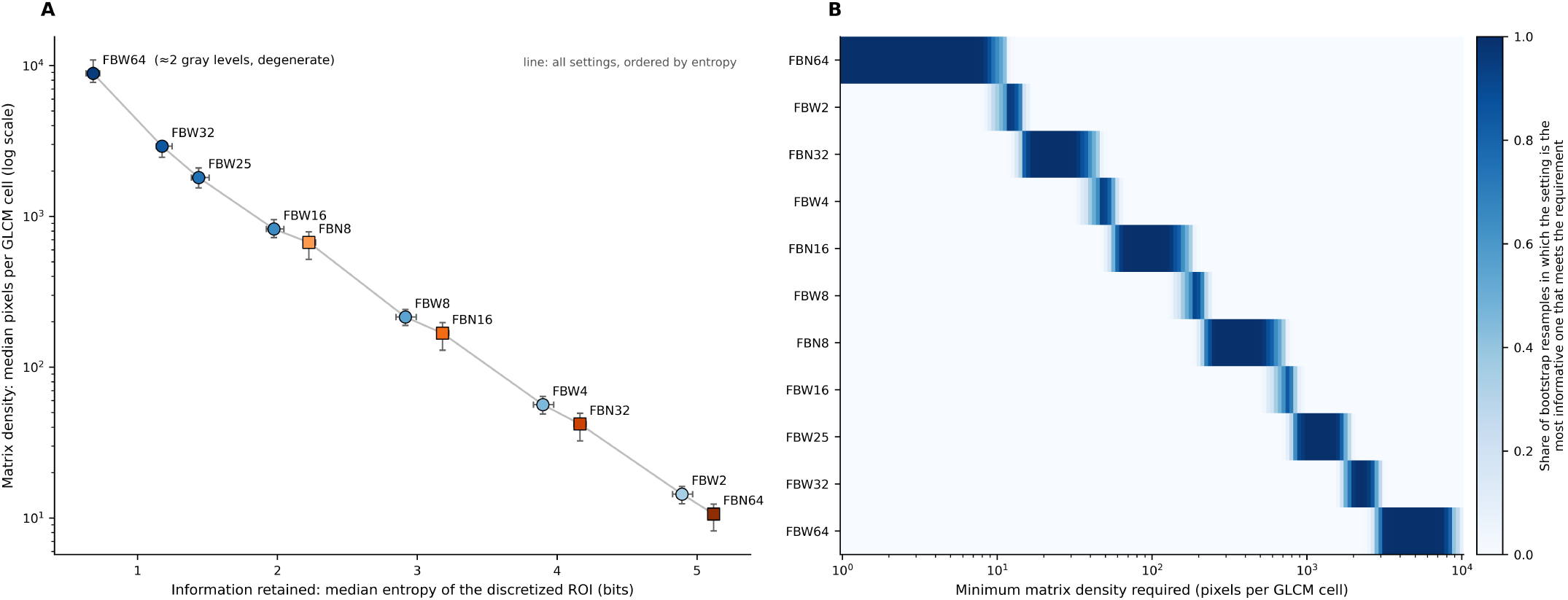
Trade-off between entropy and approximate GLCM density. (A) Median first-order entropy versus median ROI pixels per GLCM cell for each setting, with patient-level bootstrap 95% confidence intervals (1,000 resamples). Circles indicate FBW and squares FBN. Density is ROI pixel count divided by the square of occupied gray levels and is displayed on a logarithmic axis. The connecting line orders settings by entropy and does not represent a fitted model. (B) Proportion of bootstrap resamples in which each setting had the highest entropy among those meeting the minimum density threshold. Thresholds were exploratory rather than validated adequacy criteria. GLCM, gray-level co-occurrence matrix; ROI, region of interest; FBW, fixed bin width; FBN, fixed bin number.

The setting yielding the highest histogram entropy varied according to the minimum matrix density threshold. FBN 64 was selected at a threshold of 5 pixels per cell, FBW 4 at 50 pixels per cell, FBN 16 at 100, FBN 8 at 500, and FBW 25 at 1,000 pixels per cell. At a threshold of 10 pixels per cell, FBN 64 was selected in 65% of bootstrap resamples and FBW 2 the remainder. At a threshold of 20 pixels per cell, FBN 32 was selected in all resamples.

FBW 4, FBW 8, and FBN 32 had median entropy values of 3.90, 2.91, and 4.16 bits, respectively, and approximate matrix densities of 56, 215, and 42 pixels per cell. Their median ICCs were within the range observed across all settings (Table 3). FBW 2 and FBW 4 retained at least 16 occupied gray levels in nearly all ROIs, whereas no observations had fewer than 16 occupied levels under FBN 16, FBN 32, or FBN 64.

## 5 Discussion

In this study of stable CN imaged with EDI-OCT, the choice of intensity discretization settings substantially affected texture feature values and the composition of the final feature sets. In contrast, observer reproducibility was similar across settings, while slab depth had a greater influence on reproducibility than discretization. These findings indicate that discretization is an integral component of the radiomic feature definition, consistent with Image Biomarker Standardization Initiative (IBSI)[27] recommendations, rather than a neutral processing step.

Of the 93 evaluated features, 16 first-order features were independent of discretization, whereas the remaining 77 features varied across settings. This is expected because texture features depend on the distribution and spatial relationship of discretized gray levels. Although neighboring settings generally preserved the relative ranking of eyes, agreement was lower between more divergent settings and between FBW and FBN. Changes in absolute feature values do not necessarily imply equivalent changes in the ability to distinguish between lesions, but the choice of discretization can affect the information available for subsequent modeling. Discretization-induced changes in feature values and patient rankings have been reported in PET[29], while MRI studies have demonstrated effects of discretization and preprocessing on feature reproducibility[36, 37].

The choice between FBW and FBN depends on how image intensity is meant to be represented. FBW applies fixed intensity intervals, thereby retaining part of the absolute intensity information, while resulting in a variable number of gray levels depending on each ROI’s intensity range. Conversely, FBN divides each ROI’s intensity range into a predefined number of levels, effectively normalizing local intensity and yielding relative rather than absolute intensity representations. Consistent with these definitions, FBW features were more strongly associated with ROI intensity range, whereas entropy decreased with depth under FBW but remained relatively stable under FBN 64. While FBW preserves a fixed intensity scale, FBN reduces the dependence of gray-level representations on the absolute intensity range of each ROI. The weaker associations with intensity range observed under FBN are consistent with this distinction. This distinction may be relevant in OCT, where intensities are not standardized across devices. However, our study did not assess inter-device generalizability or establish whether absolute intensity differences carry clinically relevant information. Ultimately, these differences reflect the distinct intensity representations produced by absolute and relative discretization[27, 29].

Discretization strategy should be selected considering both measurement reliability and the representation of image intensities. Observer ICCs assess agreement under segmentation variation, but do not establish whether a setting preserves clinically relevant texture. This distinction was evident in our study, in which high observer ICCs coexisted with substantial intensity compression under coarse discretization. High observer agreement therefore does not, by itself, establish that a setting preserves a sufficiently detailed gray-level representation. At FBW 25, the median number of occupied levels was five, and 98% of ROI-slab observations contained fewer than eight levels. These findings indicate substantial intensity compression and argue against adopting the default bin width without examining its effects on the images under study. Discretization recommendations derived from CT or MRI cannot be transferred directly to OCT without validation. In dermatological OCT, Widaatalla et al. identified a BW 25 as a candidate setting using test-retest data[32]. Differences in tissue, acquisition, and image representation preclude directly transferring this setting choice to en face choroidal OCT.

Our results further demonstrate a trade-off between information retention, assessed using histogram entropy as a surrogate, and approximate GLCM sampling density. FBN 64 yielded slightly higher histogram entropy than FBW 2 but had a lower sampling-density proxy. This relationship is partly inherent in the definitions: finer discretization generally increases the number of occupied gray levels and histogram entropy, whereas the density proxy divides the ROI pixel count by the square of the occupied-level count. The entropy–density analysis therefore describes competing properties of the discretized representation rather than independently validating a particular setting. FBW 4 and FBN 32 occupied intermediate positions along this curve, whereas FBW 8 yielded a higher density proxy but lower histogram entropy and fewer occupied gray levels. FBW 4 and FBN 32 emerged as exploratory candidates for further study from this post hoc comparison, although no validated sampling-density threshold or evidence of clinical superiority is available to establish a universally optimal approach.

Median ICCs were high across all settings, and differences in reproducibility were small relative to variability between features. Although FBN yielded a higher proportion of features above the predefined reproducibility threshold, uncertainty in the paired comparisons did not establish superiority, and many features were close to the cutoff. Moreover, FBW 64 achieved high ICCs in deeper slabs despite typically yielding only two occupied gray levels, further illustrating that high observer agreement can coexist with a highly compressed intensity representation. Reproducibility alone is therefore insufficient to guide discretization choice. Depth had more pronounced effects: almost all features varied across slabs, and reproducibility was generally lower at 40 *μ*m than at 100-120 *μ*m. Depth should consequently be standardized and reported in retinal and choroidal studies. Technical reliability must be considered alongside the ability to characterize texture, as illustrated by phantom studies evaluating both reproducibility and texture discrimination[38].

Discretization also influenced feature redundancy and selection. The number of correlation groups decreased as the number of occupied gray levels increased, and feature sets retained after redundancy reduction overlapped less than reproducibility-eligible sets before pruning. Lower Jaccard overlap after pruning may partly reflect the selection of different representatives from similar correlation groups, rather than an equivalent change in the underlying feature information. Jaccard overlap should therefore be interpreted alongside the adjusted Rand index, which assesses agreement between the correlation-group partitions. Studies in other modalities have shown that texture indices can be redundant or strongly dependent on lesion volume, and that apparent radiomic performance can persist after spatial texture has been disrupted[39, 40].

However, these criteria do not establish a definitive optimal discretization setting. Histogram entropy does not distinguish biological signal from noise or directly quantify spatial texture, while the pixels-per-cell ratio does not measure actual GLCM occupancy. Moreover, no single setting simultaneously dominated both entropy and approximate density, leaving candidate selection dependent on exploratory density thresholds. Consequently, FBW 2 and FBN 64 remain viable options, though FBW 4 and FBN 32 offer reasonable starting points for future investigations. FBW 8 may be considered when a denser matrix is preferred, with the caveat that most ROIs contained fewer than 16 gray levels. These study-specific candidates require external validation across OCT devices, acquisition protocols, and lesion populations. Future studies should report the discretization method, its parameters, and the resulting number of gray levels.

Beyond its methodological contribution, this study provides a starting point for exploring choroidal lesions through a quantitative dimension of OCT image texture that remains largely unexplored. Characterizing radiomic patterns in stable choroidal nevi establishes a reference against which patterns associated with lesion growth or malignancy can subsequently be investigated. By showing how discretization and slab depth influence these measures, our findings support the development of standardized analytical approaches for distinguishing biological variation from variation introduced by image processing. This foundation may contribute to more reproducible and generalizable predictive models that complement conventional clinical and imaging assessment. Ultimately, the potential value of radiomic phenotyping must be evaluated against established clinical and imaging risk stratification frameworks, rather than inferred from technical stability alone[8, 22].

This study has several limitations. The cohort included only 64 eyes with CN acquired using a single OCT platform and a standardized imaging protocol; thus, findings may not generalize directly to other OCT devices, acquisition protocols, or broader populations and lesion types. The analysis was limited to two-dimensional en face EDI-OCT images and the specific depth slabs evaluated. Alternative approaches, such as three-dimensional radiomic analysis, different slab thicknesses, or custom ROI definitions, may yield different results. Observer reproducibility was assessed through repeated segmentation of fixed images, rather than repeated image acquisition, and therefore does not quantify variability arising from reacquisition, repositioning, or changes in image quality. In addition, the primary ICC analysis used feature values averaged across six slabs. Depth averaging may smooth observer differences; these ICCs therefore characterize the six-slab average and should not be assumed to represent the reproducibility of individual slabs or the TOTAL projection.

As a purely methodological investigation, this study did not evaluate clinical outcomes or diagnostic performance, such as the ability of radiomic features to discriminate CN from choroidal melanoma or predict malignant transformation. Consequently, while these results clarify feature stability, reproducibility, and redundancy across differing settings, they do not identify which discretization strategy yields the most clinically informative biomarkers. Finally, feature extraction was based on the standard PyRadiomics framework; alternative software implementations, preprocessing pipelines, or filter classes may produce different results.

## 6 Conclusions

In en face EDI-OCT radiomics of choroidal nevi, intensity discretization substantially alters feature values, relative patient rankings, feature redundancy, and downstream feature selection. Observer reproducibility and technical stability alone are insufficient to guide parameter selection. Discretization choices should also consider information retention, assessed through histogram entropy as a surrogate, gray-level representation, and approximate co-occurrence sampling, while accounting for slab depth. The choice between fixed bin width (FBW) and fixed bin number (FBN) should reflect whether fixed intensity intervals or ROI-relative discretization better suits the intended analysis. Within the evaluated acquisition and extraction pipeline, FBW 4 and FBN 32 emerged as exploratory candidate settings, with FBW 2 and FBN 64 warranting further comparison. These findings require external validation across devices, acquisition protocols, and lesion types, and the clinical utility of the extracted features remains to be established.

## Declarations

### Supplementary Materials

The Supplementary Materials contain supplementary methods, statistical analyses, and results, including Tables S1–S6 and Figures S1–S7.

## Author Contributions

M.V.F.: Conceptualization, Data Curation, Formal Analysis, Investigation, Methodology, Visualization, Writing – Original Draft, Writing – Review & Editing. J.P.N.: Formal Analysis, Methodology, Validation, Software, Visualization. D.S.: Methodology, Software, Validation. H.P.: Methodology, Software, Validation. H.F.: Methodology, Software, Validation. M.D.M.: Investigation, Validation, Writing – Review & Editing. M.L.C.: Methodology, Validation, Writing – Review & Editing.

## Ethics Approval

The study was conducted in accordance with the Declaration of Helsinki, and approved by the Ethics Committee of ULS Santa Maria, Lisbon, and ULS São José, Lisbon, with the approval numbers 94/24 and INV612 – I/2312/2025, respectively.

## Informed Consent

Informed consent was obtained from all subjects involved in the study.

## Data Availability

The article and its Supplementary Materials contain summary results only. The deidentified, feature-level dataset (radiomic feature values by eye, segmentation round, depth slab and discretization setting) and the analysis scripts are available from the corresponding author on reasonable request. The original OCT images and ROI masks are not publicly available because of patient privacy.

## Data Availability

All data produced in the present study are available upon reasonable request to the authors.

## Acknowledgments

The authors express their sincere gratitude to Dr. Filomena Pinto for her invaluable mentorship and expert guidance during the development phase of this study. They also thank Professors Mun Faria and Rita Flores for their institutional collaboration. Furthermore, the authors acknowledge the vital contributions of the ophthalmic imaging technicians—Andreia Almeida, Sandro Silva, Armandina Mota, and the wider team—without whose dedication this study would not have been possible. Additionally, thanks are due to Dr. Julio Brissos and Dr. Vasco Lomba for their assistance with data collection, and to Heidelberg Engineering for the software support provided throughout the project.

## Abbreviations

The following abbreviations are used in this manuscript:

CN: Choroidal Nevus (Nevi)
CT: Computed Tomography
EDI-OCT: Enhanced Depth Imaging Optical Coherence Tomography
FBW: Fixed Bin Width
FBN: Fixed Bin Number
GLCM: Gray-Level Co-occurrence Matrix
GLDM: Gray-Level Dependence Matrix
GLRLM: Gray-Level Run-Length Matrix
GLSZM: Gray-Level Size-Zone Matrix
HRF: Handcrafted Radiomic Features
ICC: Intraclass Correlation Coefficient
MRI: Magnetic Resonance Imaging
NGTDM: Neighboring Gray-Tone Difference Matrix
OCT: Optical Coherence Tomography
PET: Positron Emission Tomography
ROI: Region of Interest
RPE: Retinal Pigment Epithelium
SRF: Subretinal Fluid
TIFF: Tagged Image File Format

## References

[1] Yanoff, M.; Zimmerman, L.E. Histogenesis of Malignant Melanomas of the Uvea. II. Relationship of Uveal Nevi to Malignant Melanomas. Cancer 1967, 20, 493–507. 10.1002/1097-0142(1967)20:4<493::AID-CNCR2820200406>3.0.CO;2-U

[2] Arnesen, K.; Nornes, M. Malignant Melanoma of the Choroid as Related to Coexistent Benign Nevus. Acta Ophthalmol. 1975, 53, 139–152. 10.1111/j.1755-3768.1975.tb01148.x

[3] McLean, I.W.; Foster, W.D.; Zimmerman, L.E.; Gamel, J.W. Modifications of Callender’s Classification of Uveal Melanoma at the Armed Forces Institute of Pathology. Am. J. Ophthalmol. 1983, 96, 502–509. 10.1016/s0002-9394(14)77914-0

[4] Singh, A.D.; Grossniklaus, H.E. What’s in a Name? Large Choroidal Nevus, Small Choroidal Melanoma, or Indeterminate Melanocytic Tumor. Ocul. Oncol. Pathol. 2021, 7, 235–238. 10.1159/000516536

[5] Augsburger, J.J.; Corrêa, Z.M.; Trichopoulos, N.; Shaikh, A. Size Overlap between Benign Melanocytic Choroidal Nevi and Choroidal Malignant Melanomas. Invest. Ophthalmol. Vis. Sci. 2008, 49, 2823–2828. 10.1167/iovs.07-1603

[6] Damato, B.E.; Heimann, H.; Kalirai, H.; Coupland, S.E. Age, Survival Predictors, and Metastatic Death in Patients with Choroidal Melanoma Tentative Evidence of a Therapeutic Effect on Survival. JAMA Ophthalmol. 2014, 132, 605–613. 10.1001/jamaophthalmol.2014.77

[7] Damato, B. Ocular Treatment of Choroidal Melanoma in Relation to the Prevention of Metastatic Death – A Personal View. Prog. Retin. Eye Res. 2018, 66, 187–199. 10.1016/j.preteyeres.2018.03.004

[8] Shields, C.L.; Dalvin, L.A.; Ancona-Lezama, D.; Yu, M.D.; Di Nicola, M.; Williams, B.K., Jr.; Lucio-Alvarez, J.A.; Ang, S.M.; Maloney, S.; Welch, R.J.; Shields, J.A. Choroidal Nevus Imaging Features in 3,806 Cases and Risk Factors for Transformation into Melanoma in 2,355 Cases: The 2020 Taylor R. Smith and Victor T. Curtin Lecture. Retina 2019, 39, 1840–1851. 10.1097/iae.0000000000002440

[9] Shah, S.U.; Kaliki, S.; Shields, C.L.; Ferenczy, S.R.; Harmon, S.A.; Shields, J.A. Enhanced Depth Imaging Optical Coherence Tomography of Choroidal Nevus in 104 Cases. Ophthalmology 2012, 119, 1066–1072. 10.1016/j.ophtha.2011.11.001

[10] Francis, J.H.; Pang, C.E.; Abramson, D.H.; Milman, T.; Folberg, R.; Mrejen, S.; Freund, K.B. Swept-Source Optical Coherence Tomography Features of Choroidal Nevi. Am. J. Ophthalmol. 2015, 159, 169–176.e1. 10.1016/j.ajo.2014.10.011

[11] Marous, C.L.; Shields, C.L.; Yu, M.D.; Dalvin, L.A.; Ancona-Lezama, D.; Shields, J.A. Malignant Transformation of Choroidal Nevus According to Race in 3334 Consecutive Patients. Indian J. Ophthalmol. 2019, 67, 2035–2042. 10.4103/ijo.IJO_1217_19

[12] Shields, C.L.; Furuta, M.; Berman, E.L.; Zahler, J.D.; Hoberman, D.M.; Dinh, D.H.; Mashayekhi, A.; Shields, J.A. Choroidal nevus transformation into melanoma: analysis of 2514 consecutive cases. Arch Ophthalmol. 2009, 127, 981–987. 10.1001/archophthalmol.2009.151

[13] Wu, F.; Lane, A.M.; Oxenreiter, M.M.; Gragoudas, E.S.; Kim, I.K. Growth Rate of Indeterminate Choroidal Lesions Prior to Melanoma Diagnosis. Graefe’s Arch. Clin. Exp. Ophthalmol. 2023, 261, 3635–3641. 10.1007/s00417-023-06130-0

[14] Mayerhoefer, M.E.; Materka, A.; Langs, G.; Häggström, I.; Szczypiński, P.; Gibbs, P.; Cook, G. Introduction to Radiomics. J. Nucl. Med. 2020, 61, 488–495. 10.2967/jnumed.118.222893

[15] Scapicchio, C.; Gabelloni, M.; Barucci, A.; Cioni, D.; Saba, L.; Neri, E. A Deep Look into Radiomics. Radiol. Med. 2021, 126, 1296–1311. 10.1007/s11547-021-01389-x

[16] Abbasian Ardakani, A.; Bureau, N.J.; Ciaccio, E.J.; Acharya, U.R. Interpretation of radiomics features–a pictorial review. Comput. Methods Programs Biomed. 2022, 215, 106609. 10.1016/j.cmpb.2021.106609

[17] Ghalati, M.K.; Nunes, A.; Ferreira, H.; Serranho, P.; Bernardes, R. Texture analysis and its applications in biomedical imaging: A survey. IEEE Rev. Biomed. Eng. 2022, 15, 222–246. 10.1109/rbme.2021.3115703

[18] Shur, J.D.; Doran, S.J.; Kumar, S.; Ap Dafydd, D.; Downey, K.; O’connor, J.P.B.; Papanikolaou, N.; Messiou, C.; Koh, D.M.; Orton, M.R. Radiomics in oncology: A practical guide. Radiographics 2021, 41, 1717–1732. 10.1148/rg.2021210037

[19] Horvat, N.; Papanikolaou, N.; Koh, D.M. Radiomics Beyond the Hype: A Critical Evaluation Toward Oncologic Clinical Use. *Radiol*. Artif. Intell. 2024, 6. 10.1148/ryai.230437

[20] Liu, Z.; Wang, S.; Dong, D.; Wei, J.; Fang, C.; Zhou, X.; Sun, K.; Li, L.; Li, B.; Wang, M.;, et al. The Applications of Radiomics in Precision Diagnosis and Treatment of Oncology: Opportunities and Challenges. Theranostics 2019, 9, 1303–1322. 10.7150/thno.30309

[21] van Timmeren, J.E.; Cester, D.; Tanadini-Lang, S.; Alkadhi, H.; Baessler, B. Radiomics in medical imaging—“how-to” guide and critical reflection. Insights Imaging 2020, 11, 91. 10.1186/s13244-020-00887-2

[22] Aerts, H.J.W.L.; Velazquez, E.R.; Leijenaar, R.T.H.; Parmar, C.; Grossmann, P.; Carvalho, S.; Bussink, J.; Monshouwer, R.; Haibe-Kains, B.; Rietveld, D.;, et al. Decoding tumour phenotype by noninvasive imaging using a quantitative radiomics approach. Nat. Commun. 2014, 5, 4006. 10.1038/ncomms5006

[23] Magnin, C.Y.; Lauer, D.; Ammeter, M.; Gote-Schniering, J. From images to clinical insights: An educational review on radiomics in lung diseases. Breathe 2025, 21, 230225. 10.1183/20734735.0225-2023

[24] Rundo, L.; Militello, C. Image biomarkers and explainable AI: Handcrafted features versus deep learned features. Eur. Radiol. Exp. 2024, 8, 64. 10.1186/s41747-024-00465-9

[25] Zhang, H.; Zhang, H.; Jiang, M.; Li, J.; Li, J.; Zhou, H.; Song, X.; Fan, X. Radiomics in ophthalmology: A systematic review. Eur. Radiol. 2025, 35, 542–557. 10.1007/s00330-024-10911-4

[26] Gardin, I.; Grégoire, V.; Gibon, D.; Kirisli, H.; Pasquier, D.; Thariat, J.; Vera, P. Radiomics: Principles and radiotherapy applications. Crit. Rev. Oncol. Hematol. 2019, 138, 44–50. 10.1016/j.critrevonc.2019.03.015

[27] Zwanenburg, A.; Vallières, M.; Abdalah, M.A.; Aerts, H.J.W.L.; Andrearczyk, V.; Apte, A.; Ashrafinia, S.; Bakas, S.; Beukinga, R.J.; Boellaard, R.;, et al. The Image Biomarker Standardization Initiative: Standardized quantitative radiomics for high-throughput image-based phenotyping. Radiology 2020, 295, 328–338. 10.1148/radiol.2020191145

[28] Larue, R.T.H.M.; van Timmeren, J.E.; de Jong, E.E.C.; Feliciani, G.; Leijenaar, R.T.H.; Schreurs, W.M.J.; Sosef, M.N.; Raat, F.H.P.J.; van der Zande, F.H.R.; Das, M.;, et al. Influence of gray level discretization on radiomic feature stability for different CT scanners, tube currents and slice thicknesses: A comprehensive phantom study. Acta Oncol. 2017, 56, 1544–1553. 10.1080/0284186X.2017.1351624

[29] Leijenaar, R.T.H.; Nalbantov, G.; Carvalho, S.; Van Elmpt, W.J.C.; Troost, E.G.C.; Boellaard, R.; Aerts, H.J.W.L.; Gillies, R.J.; Lambin, P. The effect of SUV discretization in quantitative FDG-PET radiomics: The need for standardized methodology in tumor texture analysis. Sci. Rep. 2015, 5, 11075. 10.1038/srep11075

[30] Santinha, J.; Pinto dos Santos, D.; Laqua, F.; Visser, J.J.; Groot Lipman, K.B.W.; Dietzel, M.; Klontzas, M.E.; Cuocolo, R.; Gitto, S.; Akinci D’Antonoli, T. ESR Essentials: Radiomics—practice recommendations by the European Society of Medical Imaging Informatics. Eur. Radiol. 2025, 35, 1122–1132. 10.1007/s00330-024-11093-9

[31] Tixier, F.; Cheze Le Rest, C.; Hatt, M.; Albarghach, N.; Pradier, O.; Metges, J.-P.; Corcos, L.; Visvikis, D. Intratumor heterogeneity characterized by textural features on baseline 18F-FDG PET images predicts response to concomitant radiochemotherapy in esophageal cancer. J. Nucl. Med. 2011, 52, 369–378. 10.2967/jnumed.110.082404

[32] Widaatalla, Y.; Wolswijk, T.; Khan, M.D.; Halilaj, I.; Mosterd, K.; Woodruff, H.C.; Lambin, P. Radiomics in dermatological optical coherence tomography (OCT): Feature repeatability, reproducibility, and integration into diagnostic models in a prospective study. Cancers 2025, 17, 768. 10.3390/cancers17050768

[33] Van Griethuysen, J.J.M.; Fedorov, A.; Parmar, C.; Hosny, A.; Aucoin, N.; Narayan, V.; Beets-Tan, R.G.H.; Fillion-Robin, J.C.; Pieper, S.; Aerts, H.J.W.L. Computational radiomics system to decode the radiographic phenotype. Cancer Res. 2017, 77, e104–e107. 10.1158/0008-5472.CAN-17-0339

[34] Koo, T.K.; Li, M.Y. A guideline of selecting and reporting intraclass correlation coefficients for reliability research. J. Chirop. Med. 2016, 15, 155–163. 10.1016/j.jcm.2016.02.012

[35] Benjamini, Y.; Hochberg, Y. Controlling the false discovery rate: A practical and powerful approach to multiple testing. J. R. Stat. Soc. Ser. B 1995, 57, 289–300. 10.1111/j.2517-6161.1995.tb02031.x

[36] Duron, L.; Balvay, D.; Vande Perre, S.; Bouchouicha, A.; Savatovsky, J.; Sadik, J.-C.; Thomassin-Naggara, I.; Fournier, L.; Lecler, A. Gray-level discretization impacts reproducible MRI radiomics texture features. PLoS ONE 2019, 14, e0213459. 10.1371/journal.pone.0213459

[37] Schwier, M.; van Griethuysen, J.; Vangel, M.G.; Pieper, S.; Peled, S.; Tempany, C.M.; Aerts, H.J.W.L.; Kikinis, R.; Fennessy, F.M.; Fedorov, A. Repeatability of multiparametric prostate MRI radiomics features. Sci. Rep. 2019, 9, 9441. 10.1038/s41598-019-45766-z

[38] Bianchini, L.; Santinha, J.; Loução, N.; Figueiredo, M.; Botta, F.; Origgi, D.; Cremonesi, M.; Cassano, E.; Papanikolaou, N.; Lascialfari, A. A multicenter study on radiomic features from T2-weighted images of a customized MR pelvic phantom setting the basis for robust radiomic models in clinics. Magn. Reson. Med. 2021, 85, 1713–1726. 10.1002/mrm.28521

[39] Orlhac, F.; Soussan, M.; Maisonobe, J.-A.; Garcia, C.A.; Vanderlinden, B.; Buvat, I. Tumor texture analysis in 18F-FDG PET: Relationships between texture parameters, histogram indices, standardized uptake values, metabolic volumes, and total lesion glycolysis. J. Nucl. Med. 2014, 55, 414–422. 10.2967/jnumed.113.129858

[40] Welch, M. L.; McIntosh, C.; Haibe-Kains, B.; Milosevic, M. F.; Wee, L.; Dekker, A.; Huang, S. H.; Purdie, T. G.; O’Sullivan, B.; Aerts, H. J. W. L.; Jaffray, D. A. Vulnerabilities of Radiomic Signature Development: The Need for Safeguards. Radiother. Oncol. 2019, 130, 2–9. 10.1016/j.radonc.2018.10.027

