## Supplemental materials for "Effect of Intensity Discretization on Radiomic Features in En Face EDI-OCT of Choroidal Nevi"

### S1 Supplementary Methods

#### S1.1 RadiomicsOCT workflow

Radiomic analysis was performed using RadiomicsOCT, a custom Python application developed by the authors to support image handling, manual region-of-interest (ROI) segmentation, and radiomic feature extraction from en face OCT images. The application provides a graphical interface for TIFF image loading, optional cropping, manual ROI delineation, propagation of ROI boundaries across corresponding slabs, and adjustment of those boundaries when required. Feature calculation is implemented through PyRadiomics (version 3.1.0).

For this study, the workflow comprised image loading and grayscale conversion, delineation of the visible nevus on the TOTAL slab, propagation and adjustment of the ROI across the six numeric slabs, and batch feature extraction under the evaluated discretization settings. Within each segmentation round, the same images and masks were used for every setting, allowing discretization effects to be assessed without introducing additional segmentation differences.

#### S1.2 En face image generation, export and loading

The six numeric en face slabs were each 20 $\mu$m thick and were labeled according to the depth of their anterior boundary relative to the lower boundary of the retinal pigment epithelium/Bruch's membrane complex. The slabs labeled 20, 40, 60, 80, 100, and 120 $\mu$m therefore spanned 20–40, 40–60, 60–80, 80–100, 100–120, and 120–140 $\mu$m, respectively. The TOTAL slab extended from the anterior border of the choroidal nevus (CN) to the depth at which its signal began to fade, rather than necessarily encompassing its full anatomical thickness. En face images were generated using maximum-intensity projection. Anatomical coverage was assessed by visual inspection.

Images were exported as composite tagged image file format (TIFF) files measuring 972 $\times$ 648 pixels, with identical dimensions across eyes. The en face OCT panel measured 657 $\times$ 640 pixels, with accompanying infrared localization imagery outside this panel. Images were loaded into RadiomicsOCT, a custom Python application developed by the authors. RGB images were converted to grayscale using the weighted sum 0.2126R + 0.7152G + 0.0722B and stored as floating-point arrays. No intensity normalization or contrast stretching was applied during loading. Images were not resized within RadiomicsOCT; when cropping was performed, it selected existing pixels without interpolation. Physical pixel spacing was not assigned to the images passed to PyRadiomics, and feature extraction was performed on the exported pixel grid.

### S1.3 ROI segmentation and radiomic feature extraction

For each eye, the ROI was manually delineated around the visible CN area on the TOTAL slab and propagated to the six numeric depth slabs. Spatial correspondence was verified visually, and ROI boundaries were manually adjusted when required.

Three separate segmentation and extraction rounds were performed: observer 1, session 1; observer 1, session 2; and observer 2, session 1. Observer 1 repeated the segmentation after a 2-week interval while masked to the first segmentation. Observer 2 was masked to observer 1's segmentations. Both observers were retina-specialist ophthalmologists with extensive experience in OCT interpretation. Within each round, feature definitions and extraction parameters were held constant across discretization settings, apart from the discretization parameters being evaluated.

A total of 93 original-domain radiomic features were extracted from each ROI and slab: 18 first-order features and 75 texture features, comprising 24 gray-level co-occurrence matrix (GLCM), 16 gray-level run-length matrix (GLRLM), 16 gray-level size-zone matrix (GLSZM), 14 gray-level dependence matrix (GLDM), and 5 neighboring gray-tone difference matrix (NGTDM) features. Shape features and features derived from filtered images, including wavelet and Laplacian-of-Gaussian transforms, were excluded. Sixteen first-order features were calculated from the original grayscale intensities and were independent of discretization; first-order entropy and uniformity were calculated from the discretized histogram.

For extraction, each two-dimensional image and its binary ROI mask were represented as a single-slice volume. Two-dimensional texture calculation was explicitly enabled using `force2D=True` and `force2Ddimension=0`. Feature identifiers followed the PyRadiomics nomenclature and combined the feature family and feature name.

Texture features were calculated in two dimensions using a neighborhood distance of one pixel under the infinity norm. GLCMs were symmetric, and GLCM and GLRLM feature values were calculated separately for each of the four in-plane directions and then averaged, without distance weighting. The GLDM dependence threshold was set to zero. PyRadiomics intensity normalization, outlier removal, spatial resampling, and intensity resegmentation were disabled. The extraction configuration is summarized in Table S1.

### S1.4 Discretization settings

Eleven discretization settings were evaluated: seven fixed bin width (FBW) settings (FBW 2, 4, 8, 16, 25, 32, 64) and four fixed bin number (FBN) settings (FBN 8, 16, 32, 64).

For FBW, gray levels were grouped into bins of constant width, such that the number of resulting gray levels depended on the intensity range of each ROI. PyRadiomics positioned bin edges at integer multiples of the bin width starting from zero, with gray-level labels reindexed relative to the lowest occupied bin in each ROI. For FBN, the intensity range between the minimum and maximum ROI intensity was divided into the same number of bins for each ROI, resulting in the same nominal number of bins regardless of absolute intensity. FBW and FBN were analyzed as separate discretization families.

### S1.5 Properties of discretization settings

For each setting, the number of occupied gray levels within each ROI, the number of ROI pixels per occupied level, and the number of ROI pixels per GLCM cell were calculated. The latter was defined as the number of ROI pixels divided by the square of the number of occupied gray levels and was used as an approximate measure of co-occurrence matrix density.

**Table S1.** Radiomic feature extraction configuration.

| Component | Configuration |
|---|---|
| Extraction software | PyRadiomics, version 3.1.0. |
| Input representation | Grayscale floating-point images and binary ROI masks, represented as single-slice volumes. |
| Spatial calibration | Physical pixel spacing was not assigned to extraction images; analysis used the exported pixel grid. |
| Dimensionality | `force2D=True`; `force2Ddimension=0`. |
| Image types | Original images only; no filtered-image features. |
| Feature classes | First-order, GLCM, GLRLM, GLSZM, GLDM, and NGTDM. Shape features were excluded. |
| ROI label | `label=1`; binary masks stored as unsigned 8-bit integers. |
| Intensity normalization | Disabled: `normalize=False`. |
| Outlier removal | Disabled: `removeOutliers=None`. |
| Spatial resampling | Disabled: `resampledPixelSpacing=None`. |
| Intensity resegmentation | Disabled: `resegmentRange=None`. |
| Fixed bin width settings | `binWidth`: 2, 4, 8, 16, 25, 32, and 64. |
| Fixed bin number settings | `binCount`: 8, 16, 32, and 64; bin count took precedence over bin width. |
| Neighborhood distance | `distances=[1]` for GLCM, GLDM, and NGTDM, using the infinity norm. |
| GLCM symmetry | Symmetric matrices: `symmetricalGLCM=True`. |
| Directional aggregation | GLCM and GLRLM features calculated separately for each of four in-plane directions and then averaged; `weightingNorm=None`. |
| GLDM dependence threshold | `gldm_a=0`; dependence required equal discretized gray levels. |
| First-order intensity shift | `voxelArrayShift=0`. |

ROI, region of interest; GLCM, gray-level co-occurrence matrix; GLRLM, gray-level run-length matrix; GLSZM, gray-level size-zone matrix; GLDM, gray-level dependence matrix; NGTDM, neighboring gray-tone difference matrix. Settings describe the effective extraction configuration, including explicitly assigned parameters and unchanged PyRadiomics defaults. Intensity normalization and other preprocessing settings listed here refer to the PyRadiomics extraction stage.

The percentage of ROIs containing fewer than 8, 16, or 32 occupied gray levels was also calculated. Information retention was assessed using first-order entropy, normalized entropy, and effective number of gray levels. Normalized entropy was calculated as entropy divided by the base-2 logarithm of the number of occupied gray levels and was undefined for ROIs containing a single gray level. The effective number of gray levels was calculated as 2 raised to the median entropy.

These measures were summarized across the 448 ROIs corresponding to 64 eyes and seven slab views from session 1 of observer 1, using median with interquartile ranges (IQRs) or $5^{th}$-$95^{th}$ percentiles.

### S1.6 Effect of discretization on feature values

Analysis of discretization effects on feature values was performed using 77 discretization-dependent features from the TOTAL slab in session 1 of observer 1.

For each feature and setting, the median feature value across the 64 eyes was calculated and expressed as a ratio relative to FBW 2, which served as the reference setting. Changes were summarized for each feature family using the median absolute log2 fold-change and the percentage of features showing a change greater than two-fold. Ratios were not calculated when the median reference value was zero or when the sign of the median value changed relative to the reference.

To assess whether different settings preserved the relative ranking of eyes, Spearman correlations were calculated across the 64 eyes for each feature and every pair of settings. Correlations were summarized using the median across features and the percentage of features with a correlation ≥0.90.

The association of texture features with ROI intensity was assessed using Spearman correlation with the first-order mean and intensity range of each ROI. Median absolute correlations were calculated for each discretization setting.

### S1.7 Reproducibility analysis

Feature reproducibility was assessed using the intraclass correlation coefficient (ICC), calculated separately for each feature, discretization setting, and observer comparison. A two-way mixed-effects, single-measure, absolute-agreement model was used (ICC(A,1), using the McGraw and Wong notation).

ICC values were classified as poor (ICC <0.50), moderate (0.50 ≤ ICC <0.75), good (0.75 ≤ ICC <0.90) or excellent (ICC ≥0.90). The primary (depth-averaged) analysis used the ICC calculated on per-eye feature values averaged across the six numeric depth slabs (20–120 $\mu$m), one value per eye and segmentation round (64 eyes). ICCs were also calculated separately for each depth slab and for the TOTAL slab. The intra-observer comparison used observer 1, session 1 versus session 2; the primary inter-observer comparison used observer 1, session 1 versus observer 2, session 1. Observer 1, session 2 versus observer 2, session 1 was evaluated as an inter-observer sensitivity analysis. ROI-size agreement was evaluated using the ICC of ROI size (in pixels), the mean difference in pixels and the median absolute percentage difference, calculated as $|x - y|/[(x + y)/2] \times 100$ for each eye; these measures assessed ROI area rather than spatial overlap.

For each setting and discretization family, median ICCs and the proportion of features with ICCs ≥0.75 and ≥0.90 were calculated. Lin's concordance correlation coefficient and the mean absolute percentage difference (MAPD) between segmentation rounds were calculated as complementary measures for each feature. MAPD was defined as

$$\mathrm{MAPD} = \frac{100}{n_{\mathrm{valid}}} \sum_{i=1}^{n_{\mathrm{valid}}} \frac{|x_i - y_i|}{(|x_i| + |y_i|)/2},$$

where $x_i$ and $y_i$ are paired feature values for the same eye, and $n_{\mathrm{valid}}$ is the number of valid pairs. Pairs with a missing value or with both values equal to zero were excluded. The denominator uses absolute values to accommodate signed features. MAPD was averaged across valid eyes and was distinct from the median absolute percentage difference used for ROI-size agreement. Bland–Altman analyses were performed for depth-averaged GLCM Id, Idmn, and Idn at FBW 4 and FBN 64. Percentage differences were calculated for each eye as $100(y_i - x_i)/[(x_i + y_i)/2]$, where $x_i$ was the value from observer 1, session 1, and $y_i$ was the corresponding repeated measurement. Bias was the mean percentage difference, and limits of agreement were calculated as the bias $\pm 1.96$ times the sample standard deviation of the percentage differences.

Feature–setting pairs with ICCs <0.75 despite a small MAPD (lowest quartile of MAPD among inter-observer feature–setting pairs, ≤3.2%) were identified separately, as a low ICC may occur when between-eye variability is small despite small absolute differences between repeated measurements.

### S1.8 Variation of features with depth

Depth-dependent changes were assessed across the six numeric slabs at 20, 40, 60, 80, 100 and 120 $\mu$m using session 1 of observer 1.

For each discretization-dependent feature and setting, the Friedman test was used to assess differences across slabs, with Kendall's W as the effect-size measure. A Kendall's W $\geq$0.30 was considered a medium or larger effect.

Monotonic depth trends were assessed by calculating the Spearman correlation between depth and feature value for each eye and testing the distribution of these correlations against zero using the Wilcoxon signed-rank test. *P* values were corrected for multiple comparisons within each discretization setting using the Benjamini-Hochberg procedure with a false-discovery rate of 5%.

Depth profiles for selected features were summarized using medians and IQRs, with changes expressed relative to the 20 $\mu$m slab. Reproducibility across individual slabs was additionally assessed using the ICCs of the first-order intensity range and minimum, which were independent of discretization.

### S1.9 Redundancy among features

For each discretization setting, pairwise Spearman correlations between features were calculated using per-eye feature values averaged across the six numeric slabs, separately for each of the three segmentation rounds, using available paired observations. The TOTAL slab was not included in these averages. For each feature pair, the smallest absolute correlation across the three rounds was used as the consensus correlation. A consensus correlation was considered undefined if the corresponding correlation was undefined in any round. Undefined correlations were assigned a clustering distance of 1. Features constant in any segmentation round were assigned singleton clusters.

Features were grouped using complete-linkage hierarchical clustering with distance defined as one minus the consensus absolute correlation. At the primary threshold, every pair within a non-singleton group was required to have a consensus absolute correlation of at least 0.90. Each group was counted as one correlation group, without implying statistical independence. The primary correlation threshold was $|\rho| \geq 0.90$; thresholds of 0.80, 0.85, and 0.95 were evaluated as sensitivity analyses.

Similarity between feature groupings obtained with different discretization settings was assessed using the adjusted Rand index. The association between the number of correlation groups and the median number of occupied gray levels was assessed using Spearman correlation. This analysis was considered descriptive because the discretization settings were not independent observations.

### S1.10 Selection of reproducible features and redundancy reduction

A feature was considered reproducible within a discretization setting when its depth-averaged ICC was $\geq$0.80 for both intra-observer and primary inter-observer comparisons.

Among reproducible features, correlated features were grouped using the procedure described above. One representative feature was retained from each correlation group using a hierarchical ranking procedure. Features were ranked first by the higher minimum ICC across the intra-observer and primary inter-observer comparisons, then by the higher median ICC across those comparisons, and subsequently by the higher minimum Lin's concordance correlation coefficient. Remaining ties were resolved alphabetically by feature family and feature name. Undefined ranking values were placed last.

The number of reproducible features and retained representatives was calculated for each discretization setting and family. Uncertainty was assessed using patient-level bootstrap resampling. For each bootstrap sample, ICCs, reproducibility classification, feature correlations, and correlation groups were recalculated, and differences between settings were calculated within each resample.

The overlap between final feature sets was quantified using the Jaccard index and compared with the overlap between reproducible feature sets before redundancy reduction.

#### S1.11 Information-density trade-off

The relationship between information retention and texture-matrix density was assessed by plotting median first-order entropy against median pixels per GLCM cell for each discretization setting. Ninety-five percent confidence intervals were obtained by bootstrap resampling.

A setting was considered dominated when another setting showed both higher information retention and higher matrix density beyond bootstrap uncertainty.

Because no established threshold for acceptable matrix density was prespecified, the setting with the highest entropy was additionally identified for minimum density thresholds ranging from 1 to 10,000 pixels per cell. The proportion of bootstrap resamples in which each setting was identified as the best setting was calculated for each threshold.

### S2 Supplementary Statistical Analysis

The analysis was primarily descriptive. Discretization settings were not selected on the basis of hypothesis testing; effect sizes and confidence intervals were emphasized over *P* values where possible. Analysis-specific procedures are described in Section S1.

Confidence intervals were estimated using 1,000 patient-level bootstrap resamples with replacement and the percentile method, taking the 2.5th and 97.5th percentiles of the bootstrap distribution. A random seed of 42 was used. The same resampled eyes were used across settings and observer comparisons, preserving the paired structure. Differences between settings were calculated within each resample. All slabs, segmentation sessions, and observers belonging to each eye were kept together within each resample. No equivalence margin was prespecified. A 95% confidence interval including zero was interpreted as insufficient evidence of a difference, rather than evidence of equivalence.

Benjamini–Hochberg correction was applied separately to the Friedman tests and the monotonic-trend tests across features within each discretization setting, using a false-discovery rate of 5%. Redundancy sensitivity analyses used absolute consensus Spearman correlation thresholds of 0.80, 0.85 and 0.95 in addition to the primary threshold of 0.90.

Candidate settings were identified post hoc by their intermediate positions on the information–density curve and the absence of clear evidence of lower reproducibility. The proportion of ROIs with fewer than 16 occupied gray levels was considered separately. Minimum-density thresholds from 1 to 10,000 pixels per cell were exploratory rather than validated adequacy criteria.

Analyses were performed in Python version 3.9.23 using NumPy, pandas, SciPy and Matplotlib. The analysis code was validated against synthetic data and against the output of the radiomic extraction pipeline.

### S3 Supplementary Results

These supplementary results extend the main manuscript with ROI-size agreement, quantization and entropy summaries, observer reproducibility, depth dependence, feature redundancy and selection sensitivity. The analyses concern 64 eyes evaluated under 11 discretization settings. Detailed statistical procedures are described separately in the Supplementary Methods.

### S3.1 ROI size and agreement between segmentations

**Table S2.** ROI size and agreement across segmentation rounds.

**A. ROI size**

| Segmentations | ROI size in pixels median [IQR] |
|---|---|
| Observer 1, session 1 | 42,970 [27,042–61,216] |
| Observer 1, session 2 | 46,730 [31,914–68,634] |
| Observer 2, session 1 | 50,712 [32,074–78,243] |

**B. Pairwise agreement in ROI size**

| Comparison | ICC [95% CI] | Mean difference (pixels) | Median absolute difference (%) |
|---|---|---|---|
| Intra-observer | 0.948 [0.883–0.978] | –8,199 | 14.2 |
| Inter-observer primary | 0.948 [0.867–0.974] | –8,152 | 18.9 |
| Inter-observer sensitivity | 0.924 [0.786–0.982] | 47 | 12.7 |

All analyses include 64 eyes. Intra-observer: observer 1 session 1 minus session 2. Primary inter-observer: observer 1 session 1 minus observer 2. Sensitivity inter-observer: observer 1 session 2 minus observer 2. Mean differences follow these subtraction orders. ICC, intraclass correlation coefficient (two-way, single-measure, absolute agreement); median absolute difference is relative to the mean of each pair of ROI sizes; IQR, interquartile range; ROI, region of interest.

### S3.2 Occupied gray levels and matrix density

Across 448 ROI-slab observations per setting (64 eyes x 7 slab views), the median number of occupied gray levels fell from 54 at FBW 2 to 5 at FBW 25 and 2 at FBW 64. The percentages below 8, 16 and 32 occupied levels are reported in Table S3. FBN specifies a nominal bin count; occupied levels can be fewer when bins are empty.

**Table S3.** Quantization properties across all 11 settings.

| Setting | Occupied levels, median [P5–P95] | Pixels per level, median | GLCM density, median | <8 levels (%) | <16 levels (%) | <32 levels (%) |
|---|---|---|---|---|---|---|
| FBW 2 | 54 [36–77] | 816 | 14.4 | 0 | 0 | 2 |
| FBW 4 | 27 [18–39] | 1,610 | 56.4 | 0 | 1 | 77 |
| FBW 8 | 14 [9–20] | 3,161 | 215.1 | 1 | 71 | 100 |
| FBW 16 | 7 [5–10] | 6,151 | 824.3 | 60 | 100 | 100 |
| FBW 25 | 5 [3–7] | 9,107 | 1,810.1 | 98 | 100 | 100 |
| FBW 32 | 4 [3–5] | 11,631 | 2,917.6 | 100 | 100 | 100 |
| FBW 64 | 2 [2–3] | 19,996 | 8,868.4 | 100 | 100 | 100 |
| FBN 8 | 8 [8–8] | 5,371 | 671.4 | 0 | 100 | 100 |
| FBN 16 | 16 [16–16] | 2,686 | 167.9 | 0 | 0 | 100 |
| FBN 32 | 32 [32–32] | 1,343 | 42.0 | 0 | 0 | 1 |
| FBN 64 | 64 [63–64] | 679 | 10.6 | 0 | 0 | 0 |

Observer 1, session 1. Approximate GLCM density is ROI pixels divided by the square of occupied gray levels; it does not measure actual matrix occupancy or co-occurrence counts. Percentages describe ROI-slab observations rather than independent eyes. FBW, fixed bin width; FBN, fixed bin number; GLCM, gray-level co-occurrence matrix; P5–P95, $5^{th}$–$95^{th}$ percentiles.

### S3.3 Entropy and effective gray levels

Entropy increased by approximately 1 bit with each halving of bin width or doubling of bin number. FBW 2 and FBN 64 retained median entropies of 4.89 and 5.12 bits and normalized entropies of 0.856 and 0.854, respectively (Figure S1 and Table S4). FBW 25 retained 1.44 bits, equivalent to approximately 2.7 equally populated gray levels.

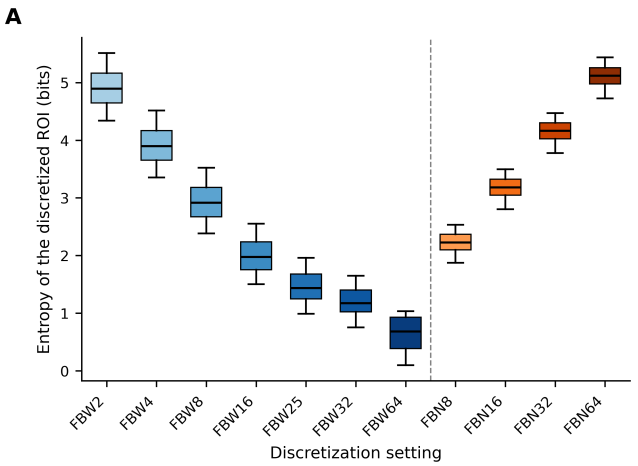


**Figure S1.** Entropy across discretization settings. (A) First-order entropy of the discretized ROI, in bits. (B) Normalized entropy, defined as entropy divided by log2 of the number of occupied gray levels. Boxes represent the interquartile range, central lines the median and whiskers the $5^{th}$–$95^{th}$ percentiles, using 448 ROI-slab observations from observer 1, session 1. Normalized entropy is undefined for observations containing a single occupied level. Dashed lines separate FBW and FBN. ROI, region of interest; FBW, fixed bin width; FBN, fixed bin number.

**Table S4.** Entropy and effective gray levels.

| Setting | Entropy bits median [IQR] | Normalized entropy median [IQR] | Effective gray levels |
|---|---|---|---|
| FBW 2 | 4.89 [4.65–5.16] | 0.856 [0.829–0.880] | 29.6 |
| FBW 4 | 3.90 [3.65–4.17] | 0.823 [0.792–0.852] | 14.9 |
| FBW 8 | 2.91 [2.67–3.18] | 0.777 [0.736–0.812] | 7.5 |
| FBW 16 | 1.98 [1.76–2.24] | 0.703 [0.659–0.754] | 3.9 |
| FBW 25 | 1.44 [1.25–1.68] | 0.654 [0.595–0.707] | 2.7 |
| FBW 32 | 1.17 [1.02–1.40] | 0.628 [0.558–0.688] | 2.2 |
| FBW 64 | 0.68 [0.39–0.93] | 0.582 [0.367–0.856] | 1.6 |
| FBN 8 | 2.23 [2.10–2.37] | 0.742 [0.700–0.790] | 4.7 |
| FBN 16 | 3.18 [3.05–3.32] | 0.795 [0.762–0.831] | 9.1 |
| FBN 32 | 4.16 [4.02–4.30] | 0.833 [0.805–0.861] | 17.9 |
| FBN 64 | 5.12 [4.97–5.26] | 0.854 [0.830–0.878] | 34.8 |

Summaries use 448 ROI-slab observations per setting from observer 1, session 1. Normalized entropy is entropy divided by log2 of occupied gray levels and is undefined for a single occupied level. Effective gray levels are 2 raised to the median entropy. IQR, interquartile range.

### S3.4 Feature values and associations with ROI intensity

At FBW 25, typical changes relative to FBW 2 were approximately 8-fold for GLCM, 9-fold for GLDM, 32-fold for GLRLM, 64-fold for GLSZM and 160-fold for NGTDM features. Between 67% and 100% of features in each family changed by more than two-fold. At FBN 64 typical changes were 1.2–1.9-fold, although 25–44% of texture features still changed by more than two-fold. These analyses used the TOTAL slab from observer 1, session 1.

Rank agreement was lower between FBW 2 and FBW 25 (median Spearman $\rho$, 0.82; 17% of features with $\rho$ ≥0.90) than between FBW 2 and FBW 4 (0.99; 90%). Cross-family agreement was lower: FBW 2 versus FBN 64 yielded a median $\rho$ of 0.59 (19% with $\rho$ ≥0.90), and FBW 25 versus FBN 64 yielded 0.46 (8%).

Median absolute correlations with ROI intensity range were 0.51–0.59 for FBW 2–25 and 0.41 for FBW 32, compared with 0.21–0.25 for FBN settings. Correlations with mean ROI intensity were weaker (median absolute $\rho$, 0.09–0.17), apart from FBW 64, whose approximately two occupied levels limited interpretation.

### S3.5 Observer reproducibility and absolute agreement

For the primary depth-summary analysis, within-setting dispersion of ICCs across features was greater than the differences in median ICC between settings. For intra-observer reproducibility, the median ICC was 0.91 [95% CI, 0.85–0.94] at FBW 2 and 0.88 [0.81–0.94] at FBN 64. None of the 16 prespecified paired comparisons between settings (eight per observer comparison) provided clear evidence of a difference (Table S5). With 64 eyes, differences of up to approximately 0.08 in median ICC could not be excluded.

**Table S5.** Paired differences in median depth-averaged ICC between discretization settings.

| | Difference in median ICC [95% CI] | |
|---|---|---|
| **Contrast** | **Intra-observer** | **Primary inter-observer** |
| FBW 4 − FBW 2 | −0.008 [−0.028, 0.008] | −0.018 [−0.035, 0.010] |
| FBW 25 − FBW 2 | −0.018 [−0.053, 0.001] | −0.008 [−0.031, 0.021] |
| FBN 64 − FBW 2 | −0.023 [−0.073, 0.020] | 0.008 [−0.023, 0.063] |
| FBN 64 − FBW 4 | −0.015 [−0.062, 0.034] | 0.026 [−0.017, 0.081] |
| FBW 25 − FBN 64 | 0.005 [−0.061, 0.056] | −0.016 [−0.062, 0.013] |
| FBN 32 − FBN 64 | 0.002 [−0.013, 0.014] | −0.004 [−0.021, 0.003] |
| FBN 16 − FBN 64 | −0.008 [−0.024, 0.013] | −0.002 [−0.020, 0.007] |
| FBN 8 − FBN 64 | 0.012 [−0.014, 0.029] | 0.000 [−0.018, 0.013] |

Medians summarize the 77 discretization-dependent features. Feature-level ICCs were calculated using per-eye values averaged across the six numeric slabs. Differences follow the subtraction order shown. Confidence intervals were estimated using 1,000 paired patient-level bootstrap resamples and the percentile method. All intervals include zero; this does not establish equivalence. ICC, intraclass correlation coefficient; CI, confidence interval; FBW, fixed bin width; FBN, fixed bin number.

FBN settings had 92–95% of features with intra-observer ICC ≥0.75 and 38–47% with ICC ≥ 0.90; corresponding FBW estimates were 81–86% and 43–52%. Primary inter-observer estimates reached 99% with ICC ≥0.75 under FBN. Confidence intervals overlapped widely, so these point estimates do not establish a difference between discretization families.

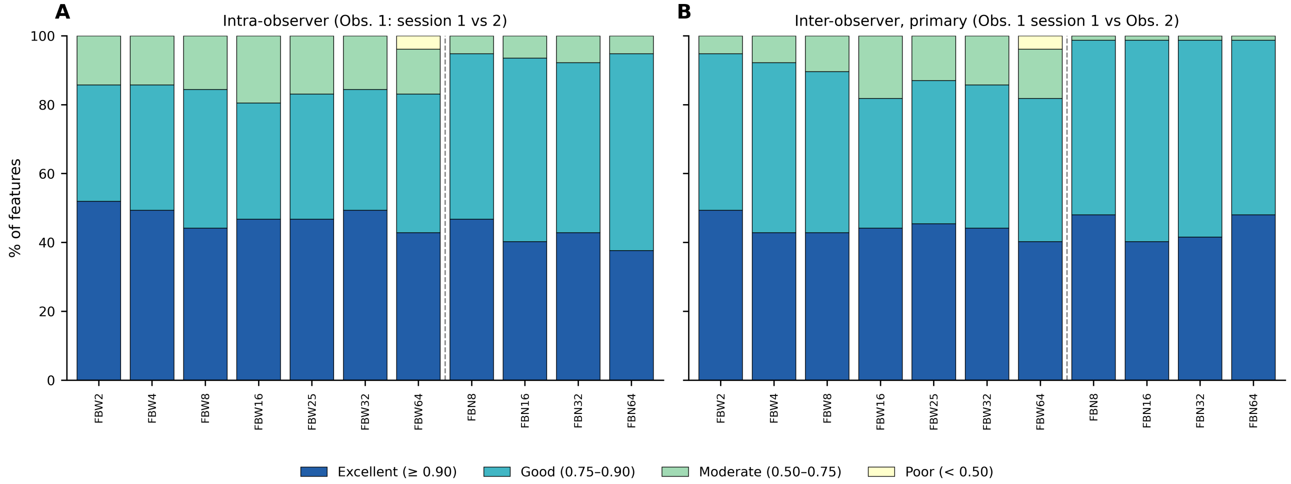


**Figure S2.** Distribution of reproducibility categories by discretization setting. (A) Intra-observer and (B) primary inter-observer comparisons, using the primary depth-summary ICCs of the 77 discretization-dependent features. Categories are poor (ICC <0.50), moderate (0.50 ≤ ICC <0.75), good (0.75 ≤ ICC <0.90) and excellent (ICC ≥0.90). Dashed lines separate FBW and FBN. ICC, intraclass correlation coefficient; FBW, fixed bin width; FBN, fixed bin number.

### S3.6 TOTAL slab and complementary agreement measures

Median ICCs for the TOTAL slab ranged from 0.76 to 0.88. Point estimates were lower under FBN (0.76–0.80) than under FBW (0.81–0.88), but confidence intervals for paired differences between settings included zero. For intra-observer reproducibility, the FBN 64 minus FBW 2 difference was –0.087 [95% CI, –0.209 to 0.002].

ICC and Lin's concordance correlation coefficient yielded closely similar results (correlation, 0.99999; maximum absolute difference, 0.004). In 12 of 847 feature–setting combinations, inter-observer ICC was below 0.75 despite a mean absolute percentage difference of 3.2% or less. These combinations involved the GLCM features maximal correlation coefficient (`MCC`), Correlation, and informational measure of correlation 2 (`Imc2`), which showed little between-eye variation.

Bland–Altman analyses of GLCM Id, Idmn, and Idn illustrated differences in the scale of observer disagreement (Figure S3). For the primary inter-observer comparison, Id showed limits of agreement of $-2.61\%$ to $+0.79\%$ at FBW 4 and $-3.37\%$ to $+5.96\%$ at FBN 64. Corresponding limits were substantially narrower for Idmn ($-0.0198\%$ to $+0.0365\%$ and $-0.0191\%$ to $+0.0320\%$) and Idn ($-0.180\%$ to $+0.313\%$ and $-0.193\%$ to $+0.337\%$), respectively. These analyses used six-slab mean feature values and describe agreement between segmentation rounds within each setting; they do not establish invariance across discretization settings.

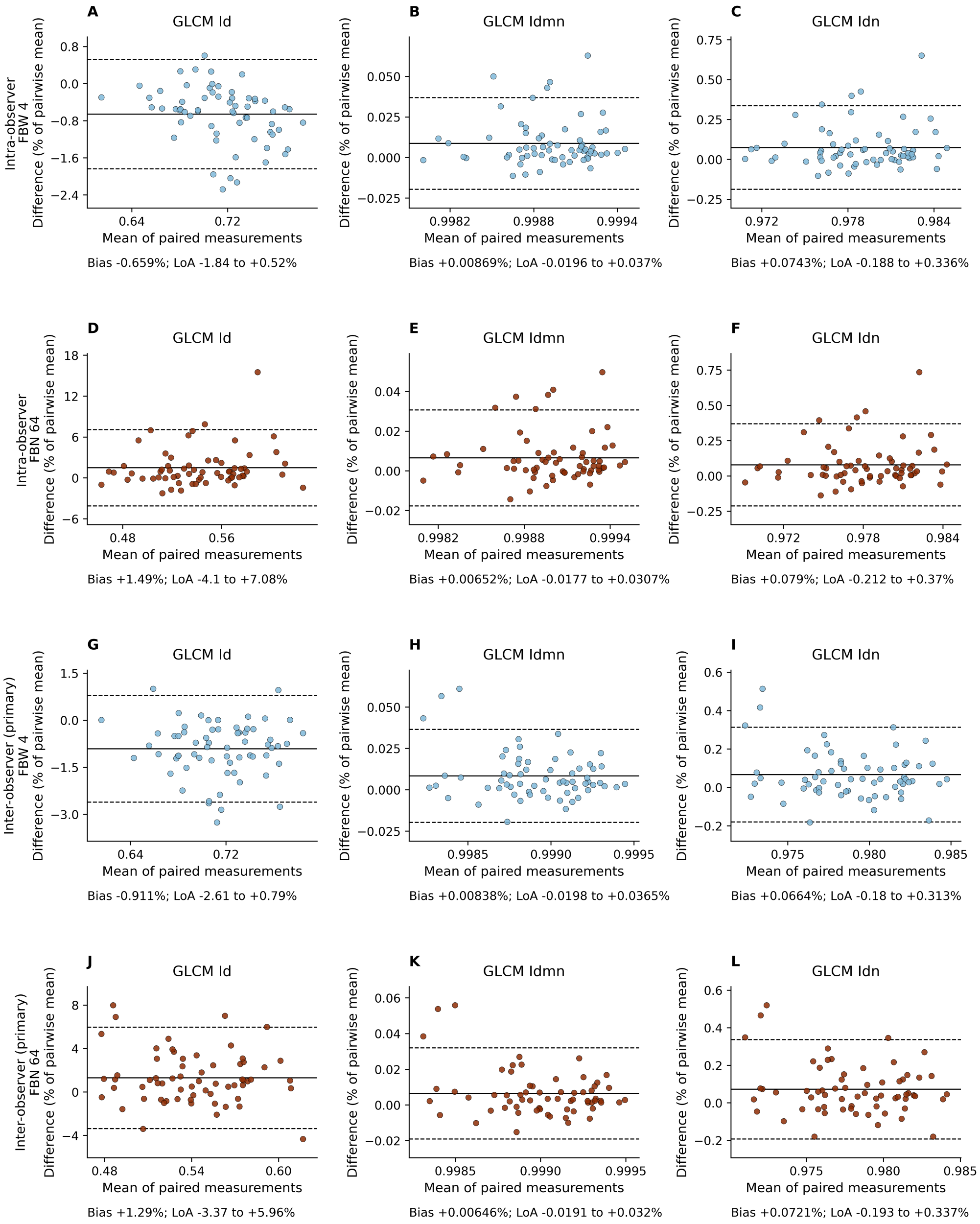


**Figure S3.** Bland–Altman plots for GLCM Id, Idmn, and Idn at FBW 4 and FBN 64. Analyses used per-eye feature values averaged across the six numeric slabs. Columns show Id, Idmn, and Idn, respectively. Rows show the intra-observer comparison at FBW 4 (A–C) and FBN 64 (D–F), followed by the primary inter-observer comparison at FBW 4 (G–I) and FBN 64 (J–L). Differences were calculated as observer 1, session 2 minus session 1 for the intra-observer comparison, and observer 2, session 1 minus observer 1, session 1 for the inter-observer comparison. Horizontal axes show the mean of the paired measurements. Vertical axes show their difference as a percentage of the pairwise mean. Solid lines indicate mean differences; dashed lines indicate limits of agreement, calculated as the mean difference $\pm 1.96$ times the sample standard deviation of the percentage differences. Axis scales differ between panels. GLCM, gray-level co-occurrence matrix; Id, inverse difference; Idmn, inverse difference moment normalized; Idn, inverse difference normalized; FBW, fixed bin width; FBN, fixed bin number.

### S3.7 Depth dependence

Mean ROI intensity decreased by approximately 4% at 40 $\mu$m and 43% at 120 $\mu$m relative to the 20 $\mu$m slab. Entropy decreased under FBW, by approximately 5–7% at FBW 2 and FBW 4 and 15% at FBW 25, whereas changes under FBN 64 were smaller (–3% to –0.2%). At 120 $\mu$m, Contrast decreased by 46% at FBW 2, 42% at FBW 4, 28% at FBW 25 and 30% at FBN 64. NGTDM Coarseness did not show a consistent monotonic profile (Figure S4).

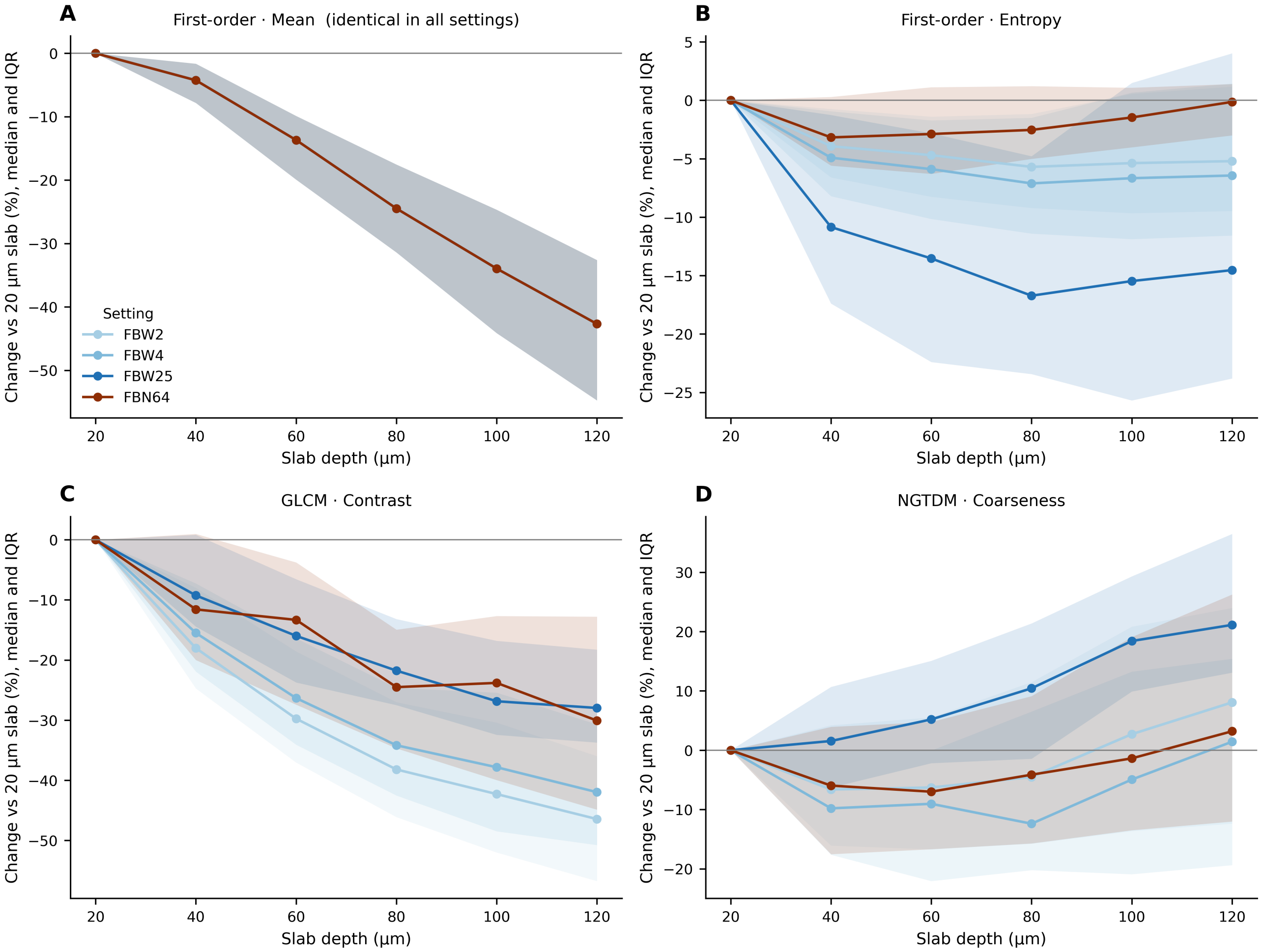


**Figure S4.** Depth profiles of selected features. Percentage changes relative to the 20 $\mu$m slab are shown for (A) first-order mean intensity, (B) first-order entropy, (C) GLCM Contrast and (D) NGTDM Coarseness under FBW 2, FBW 4, FBW 25 and FBN 64. Lines show the median and shaded bands the interquartile range across 64 eyes, using observer 1, session 1. Mean intensity is independent of discretization, so its profiles coincide. GLCM, gray-level co-occurrence matrix; NGTDM, neighboring gray-tone difference matrix; FBW, fixed bin width; FBN, fixed bin number.

### S3.8 Magnitude and monotonicity of depth variation

After correction for multiple testing, 94–100% of discretization-dependent features varied significantly across the six numeric slabs. Median Kendall's W was 0.31 at FBW 2 and FBW 4, 0.20 at FBW 32 and 0.09 at FBW 64; FBN values ranged from 0.24 to 0.28 (Figure S5). The upper quartile reached 0.66 at FBW 2 and 0.33 at FBN 64.

Features with W ≥0.30 accounted for 52–57% at FBW 2 and FBW 4 and 36–42% at FBN 16–64. Significant monotonic trends occurred in 90–92% of features under FBW 2–25 and 78–82% under FBN. These results describe depth dependence and do not establish which gradients are clinically informative.

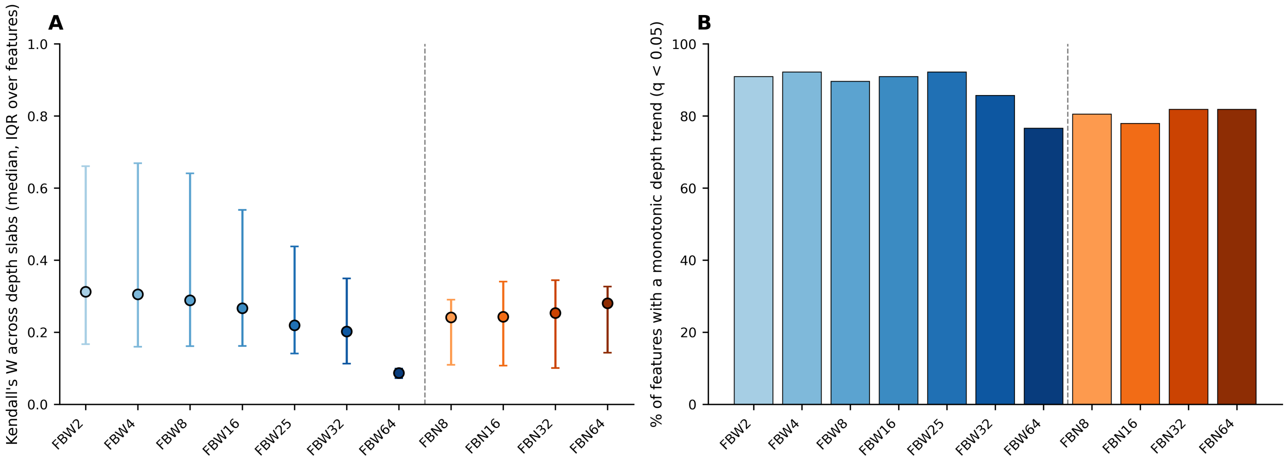


**Figure S5.** Magnitude and monotonicity of depth-related feature variation. (A) Median Kendall's W across the 77 discretization-dependent features, with interquartile ranges. These bars describe variability across features, not confidence intervals. (B) Percentage of features with a significant monotonic depth trend after Benjamini–Hochberg correction (q <0.05). Analyses use six numeric depth slabs from observer 1, session 1. Dashed lines separate FBW and FBN. FBW, fixed bin width; FBN, fixed bin number.

### S3.9 Feature redundancy and grouping similarity

At the primary absolute-correlation threshold of 0.90, group counts among the 77 discretization-dependent features ranged from 28 at FBN 64 to 41 at FBW 64 (Table S6). The correlation between group count and occupied gray levels across the 11 settings was −0.92; this was a descriptive association across settings.

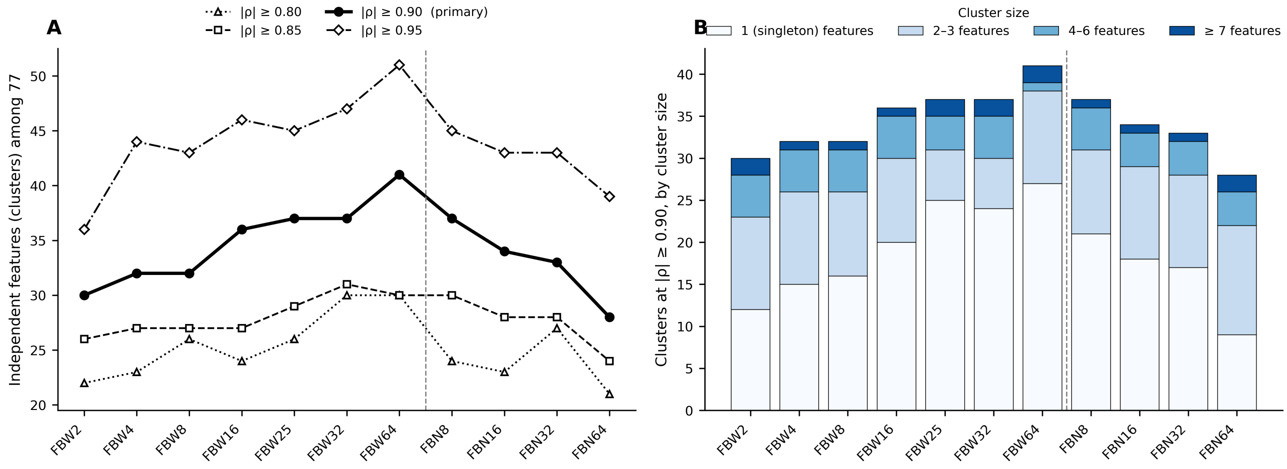


**Figure S6.** Feature correlation groups across discretization settings. (A) Number of groups among the 77 discretization-dependent features at consensus absolute Spearman correlation thresholds of 0.80, 0.85, 0.90 and 0.95. The primary threshold is 0.90. (B) Number of groups at the primary threshold, classified by group size. Consensus correlations use the smallest absolute correlation across the three segmentation rounds; complete-linkage grouping requires all feature pairs within each group to meet the threshold. Group counts are distinct from the number of features retained after reproducibility screening. Dashed lines separate FBW and FBN. FBW, fixed bin width; FBN, fixed bin number.

**Table S6.** Correlation groups among 77 discretization-dependent features.

| Setting | Number of correlation groups |
|---|---|
| FBW 2 | 30 |
| FBW 4 | 32 |
| FBW 8 | 32 |
| FBW 16 | 36 |
| FBW 25 | 37 |
| FBW 32 | 37 |
| FBW 64 | 41 |
| FBN 8 | 37 |
| FBN 16 | 34 |
| FBN 32 | 33 |
| FBN 64 | 28 |

The primary threshold was consensus $|\rho| \geq 0.90$ across the three segmentation rounds, using complete-linkage groups. Counts include all 77 discretization-dependent features and differ from final feature counts after reproducibility screening.

### S3.10 Grouping sensitivity and final feature sets

Across discretization families, grouping similarity was highest for settings with comparable numbers of occupied levels: adjusted Rand indices were 0.83 for FBN 16 versus FBW 8, 0.71 for FBN 32 versus FBW 4 and 0.67 for FBN 64 versus FBW 2. FBW 2 versus FBW 4 yielded 0.51. Comparisons with markedly different occupied-level counts yielded indices of 0.22–0.37. Thresholds of 0.80 or 0.85 produced approximately 21–31 groups, whereas 0.95 produced 36–51; the overall pattern persisted.

Bootstrap intervals for the number of reproducible features were wide and overlapped across settings. For example, FBN 64 minus FBW 2 yielded +15 features [95% CI, –11 to +26], and FBN 64 minus FBW 4 yielded +13 [–9 to +27]. After redundancy reduction, the FBN 64 minus FBW 2 difference was +4 features [–2 to +8]. The point estimates therefore do not establish a difference in the number of retained features.

Final sets contained 5–7 discretization-independent first-order statistics and texture features, most frequently from the GLSZM family. For discretization-dependent features, the Jaccard index between FBW 2 and FBW 4 decreased from 0.89 before redundancy reduction to 0.59 afterwards; for FBN 32 versus FBN 64, it decreased from 0.91 to 0.58. Within this feature population, final-set overlap was 0.52 for FBW 2 versus FBN 64, 0.40 for FBW 25 versus FBW 2, and 0.28 for FBW 25 versus FBN 64.

Pairwise overlap between reproducibility-eligible discretization-dependent feature sets, overlap between final representative sets, and feature-selection frequencies across settings are shown in Figure S7.

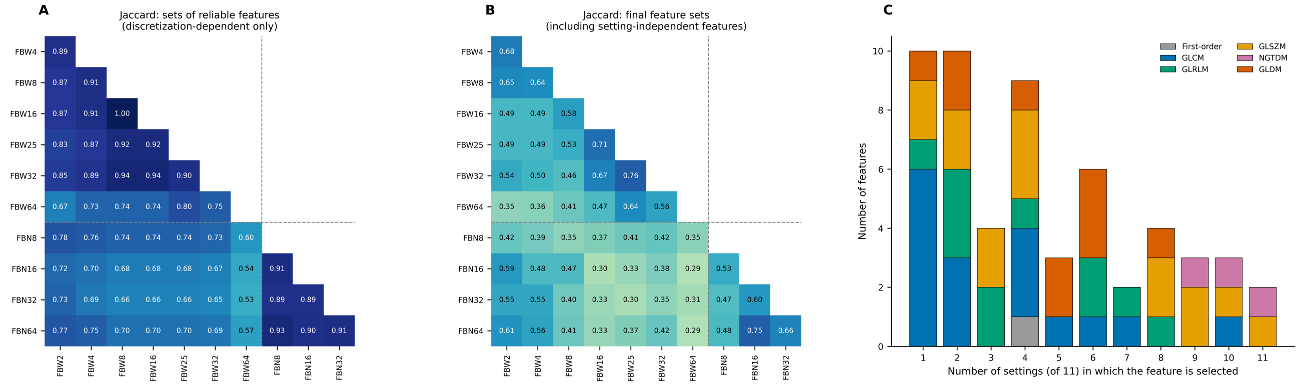

**Figure S7.** Feature-set overlap and selection frequency across discretization settings. (A) Pairwise Jaccard indices between reproducibility-eligible feature sets, restricted to the 77 discretization-dependent features. (B) Pairwise Jaccard indices between final representative feature sets after reproducibility screening and redundancy reduction, including discretization-independent features. Panels A and B describe different feature populations and should not be interpreted as a direct before–after comparison. Dashed lines separate FBW and FBN settings. (C) Number of features retained in exactly 1 to 11 discretization settings, with colors indicating feature families. Reproducibility required ICC $\geq 0.80$ in both intra-observer and primary inter-observer comparisons; redundancy reduction retained one representative per correlation group using a consensus absolute Spearman correlation threshold of 0.90. ICC, intraclass correlation coefficient; FBW, fixed bin width; FBN, fixed bin number; GLCM, gray-level co-occurrence matrix; GLRLM, gray-level run-length matrix; GLSZM, gray-level size-zone matrix; GLDM, gray-level dependence matrix; NGTDM, neighboring gray-tone difference matrix.

### S3.11 Exploratory density thresholds

No discretization setting was dominated simultaneously in entropy and approximate GLCM density. The setting with the highest entropy subject to a minimum density varied with that threshold: FBN 64 at 5 pixels per cell, FBN 32 at 20, FBW 4 at 50, FBN 16 at 100, FBN 8 at 500 and FBW 25 at 1,000. These thresholds were exploratory and were not validated adequacy criteria.

At 10 pixels per cell, FBN 64 provided the highest entropy in 65% of patient-level bootstrap resamples and FBW 2 in the remaining 35%. FBN 32 was selected in all resamples at 20 pixels per cell, and FBW 4 in 91% at 50. The spacing of tested settings influences the threshold ranges over which a setting is selected. These findings support comparison of candidate settings rather than a universal optimum.
